# Is Standard Personal Protective Equipment Effective Enough To Prevent COVID-19 Transmission During Aerosol Generating Dental, Oral and Maxillofacial Procedures ? A Systematic Review

**DOI:** 10.1101/2020.11.20.20235333

**Authors:** Essam Ahmed Al-Moraissi, Marwan Mohamed Abood, Nasser A. Alasseri, Frank Günther, Andreas Neff

**Affiliations:** Assisant professor, Dept. of Oral and Maxillofacial Surgery, Faculty of Dentistry, Thamar University, Thamar, Yemen; Consultant, Dept. of Oral and Maxillofacial Surgery, Regional Dental Center, Qassim, Dept. of Oral and Maxillofacial Surgery Ministry of Health, Saadia Arabia; Senior register, Department of Periodontics, Regional dental center, Qassim, Dept. of Oral and Maxillofacial Surgery Ministry of health, Saadia Arabia; Consultant, Department of Oral & Maxillofacial Surgery, Prince Sultan Military Medical City, Riyadh, Saudi Arabia; Professor,Medical Microbiology and Hygiene, Marburg University Hospital, Hans-Meerwein-Strasse 2, D-35043, Marburg, Germany; Professor, Dept. of Oral and Maxillofacial Surgery, University Hospital Marburg UKGM GmbH, Marburg, Germany

## Abstract

A systematic review was performed to answer the following questions: 1) Do dental, oral and maxillofacial (OMF) surgical procedures generate bioaerosols (and if so, which ones), which can result in transmission of COVID-19?; 2) Are aerosolized airborne droplets (and to which extent is splatter) in dental and OMF procedures infective?; 3) Is enhanced personal protective equipment (PPE) an essential to prevent spreading of COVID-19 during dental and OMF aerosol generating procedures (AGPs)? Authors performed a systematic review to retrieve all pertinent literature that assessed effectiveness of surgical mask vs respirators for protecting dental health care workers during dental and OMF AGPs surgical procedures. Additionally, studies which assessed potential aerosolization during dental, OMF and orthopaedic surgeries were retrieved. There is moderate evidence showing that ultrasonic scaling and bone drilling using high speed rotary instruments produces respirable aerosols. Additionally, there is very weak/inconclusive evidence to support the creation of infectious aerosols during dental procedures. According to available very weak/inconclusive evidence, transmission of SARS-CoV-2 via infective aerosol during AGPS, so far, must remain speculative and controversial. As, however, this is a probable opportunistic way of transmission which at least cannot be sufficiently excluded and therefore should not be dismissed out of hand prematurely, proper and equally important properly applied protective equipment (i.e., N95 respirators or FFP-2 masksv or above regarding mouth and nose protection) should always be used during AGPs.

## Introduction

Coronavirus disease 2019 (Group) or severe acute respiratory syndrome 2 (SARS CoV 2) is a considerably contagious disease that has rapidly spread across the globe, making it a global public threat(Ge et al., 2020). Although the full route of transmission is incompletely understood, COVID-19 is predominantly transmitted by respiratory droplets and fomites via aerosols inhalation, coughing or sneezing through direct contact of symptomatic (Health and Safety Executive, 2008; Chen et al., 2020), asymptomatic (Backer et al., 2020; Guan et al., 2020; Huang et al., 2020; Rothe et al., 2020; Wu et al., 2020) or presymptomatic infected patients, or indirectly, by touching virus contaminated surfaces and consequently contacting mucus membrane of mouth, eyes and nose. (England., 2020; Lu et al., 2020; Peng et al., 2020; van Doremalen et al., 2020). Additionally, airborne transmission of small respiratory particles (≤ 5μm) has been suggested as potential transmission route for COVID-19. (Tran et al., 2012; Ai et al., 2019; Peng et al., 2020; Santarpia et al., 2020b; Schwartz et al., 2020; Thamboo et al., 2020; Wax et al., 2020)

The prevalence rate of COVID-19 among personal health care providers has been reported to be 3.8 % and 20% in Wuhan, China (Wang et al., 2020) and Italy (Remuzzi et al., 2020), respectively. From January to April 2020, more than 150 health care workers in Italy and over 100 National Health Service workers in the UK lost their lives in the COVID-19 pandemic outbreak (Warnakulasuriya, 2020). In addition, since 23 April 2020, 116 health care workers have died in Italy due to COVID-19, among them 12 dentists (Peditto et al., 2020)

Oral mucous membrane has been reported to have a high affinity for angiotensin converting enzymes receptor 2 (ACE2) which is responsible for the entrance of the virus into human cells, then starting its replications(Xu et al., 2020). So, saliva may contain more viral load (Liu et al., 2011).

Therefore, oral and maxillofacial surgeons (OMS) face a substantial risk of exposure to SARS-CoV-2 and thus COVID-19, as their actual field of work lies in close proximity to both oral cavity and naso-/oropharynx. Similar to the influenza virus, the primary SARS-CoV-2 viral load is considered as an indicator for severity of infection. Thus, professional health care workers including OMS among them in the front ranks are at a significant risk of acquiring the virus due to their exposure to higher viral load (Paulo et al., 2010; Burdorf et al., 2020; Zimmermann et al., 2020).

Aerosols refers to liquid and solid particles (≤ 5 μm) which dehydrate and retain in air. Although range information is rather critical for aerosols and may vary greatly depending on literature, it can be assumed that aerosols can persist in the air for a very long time and therefore can spread completely in enclosed spaces. Aerosols are estimated to travel between e.g. up to 4.5 m (Loh et al., 2020) and 27 feet (around 8 m), or room scale(Morawska et al., 2020), respectively. They remain suspended in the air for hours prior to falling on the ground or entering the respiratory system, whereas, droplets are described as larger entities (>5μm) that rapidly drop to the ground by force of gravity, typically 3-6 feet (1-2m) of the source person(Klompas et al., 2020); droplets and splatter is a mixture of air, water, saliva and/or solid particles; when greater than 30-50 μm they are usually visible to the naked eye (Micik et al., 1969; Micik et al., 1971; Miller et al., 1978; Harrel, 2004).

Respirators such as filtering face pieces FFP2 masks are tested and classified according to their retention capacity against particles of 0.5 µm. Analogously, N95 and N100 protection masks are at least 94-95% and 99%, respectively, effective for particles tested according to the standards of the National Institute for Occupational Safety and Health (NIOSH), which increases to 99.5% or higher for particles measuring 0.75 micron or bigger (Qian et al., 1998). In contrast, medical/surgical facemasks were observed to offer very little protection for particle sizes 10–80 nm (Balazy et al., 2006). Although there have been no studies comparing the effectiveness of medical/surgical masks vs. N95/100 respirators and FFP2/3 masks in reducing the spread of COVID-19, there is some evidence supporting superiority of the N95 respirator over surgical masks against viral aerosolization (Seto et al., 2003; Teleman et al., 2004; Wen et al., 2013; Garcia Godoy et al., 2020). Based e.g. on influenza virus data (diameter 0.08-0.12 microns), however, recent systematic reviews and meta-analyses of randomized clinical trials concluded that N95 respirators did not reduce laboratory confirmed respiratory viral infection among health care workers when compared to surgical masks (Loeb et al., 2009; Radonovich et al., 2019; Bartoszko et al., 2020; Long et al., 2020)

All dental surgical procedures performed with high-speed rotating handpiece, ultrasonic scaler, water air syringe are aerosol generating procedures (AGPs) which are mostly contaminated with blood, bacteria, viruses and fungi (Bentley et al., 1994; Harrel et al., 2004; Sacchetti et al., 2006; Ishihama et al., 2008; Szymanska et al., 2013; Al-Eid et al., 2018). The same may apply for surgical procedures such as bone drilling under irrigation and exposure to blood and saliva contaminated droplets and splatters during e.g. coughing, and tracheobronchial aerosols, which may bear a high virus load.

Considering choice of personal protective equipment (PPE) health care professionals need to take into account that also pre-symptomatic and asymptomatic patients are suited to transmit COVID-19(Korth et al., 2020; Schmidt et al., 2020). Several societies and national associations (such as American Dental Association, U.S. Occupational Safety and Health Administration, American Society of Dentist Anesthesiologists, British Association of Oral and Maxillofacial Surgeons, The French Society of Stomatology, Maxillo-Facial Surgery and Oral Surgery, British Society of Oral Surgeons etc.) recommend that a surgical (or medical) mask is not sufficient. Instead, N95 or higher level of respirators or /FFP2 or 3 masks are mandatory, especially when conducting AGPs for suspected and confirmed COVID-19 patients. (BAOMS, 2020; French Society of Stomatology et al., 2020a; Occupational safety and health administrations, 2020; The American Society of Dentist Anesthesiologists, 2020) *Covid-19 Dental Services Evidence Review (Group) Working Group Version 1.3 including 16 countries* (Group, 2020)

Correspondingly, the U.S Centers for Disease Control and Prevention (CDC), and European Centre for Disease and Prevention (ECDC) recommend the N95 respirator/FFP2 mask for non-aerosol generating routine care of patients with COVID-19 (Centers for Disease Control and Prevention, 2020b; European Centre for Disease Prevention and Control, 2020), whereas the World Health Organization, Public Health England and the Public Health Agency of Canada recommend the use of medical/surgical masks for this indication. (Public Health Agency of Canada, 2020; Public Health England, 2020a; World Health Organization, 2020)

Currently, the coronavirus is not considered to be an airborne virus, so airborne precautions are not routinely necessary (Douedi S, 2020; Public Health England, 2020b). The World Health Organization has emphasized that further studies are required to confirm whether COVID-19 is spread via aerosols (Organization, 2020). At present, airborne transmission of COVID-19 from individual to individual for long distance is considered improbable (Centers for Disease Control and Prevention, 2020a). However, close proximity of OMS to their patients during AGPs makes them susceptible to contract SARS-CoV-2 and thus COVID-19.

Other studies reported the presence of COVID-19 in the air sample of hospital rooms of COVID-19 patients (H.; Santarpia et al., 2020a), so air conditioning and ventilation and respective ventilation numbers (i.e. air exchange rate and air displacement) in surgeries with AGP might play a crucial role in indoor transmission and need to be considered, as well. In this context, the National Health Committee of China has stated that “Exposure to high-concentration contaminated aerosols in a relatively closed space for a long time, may lead to transmission of COVID-19 via aerosols”(Li et al., 2004)

Standard PPE consist of a surgical mask, face shield (usually with inferior opening) or protection goggles, gown, surgical cap, and gloves (Panesar et al., 2020). Full or enhanced PPE include a surgical mask, face shield or protection goggles, disposable long-sleeved gown (waterproof), surgical cap, and gloves in addition to respirators such as N95/FFP2, filtering facepieces respirators or powered air purifying respirators

Currently, there is lack of scientific evidence regarding whether aerosolized airborne droplets generated during dental and maxillofacial surgical procedures are infective in COVID-19, and which type of personal protective equipment is effective enough to prevent COVID-19 transmission during aerosol generating dental and maxillofacial procedures?

Thus, there is urgent need to conduct a systematic review to identify whether there is scientific evidence supporting that dental/OMS surgical procedures are AGPs and whether bioaerosols produced at dental and maxillofacial surgical practices can transmit COVID-19. To the best of the authors’ knowledge, there is no scientific evidence discussing which OMF surgical procedures are truly aerosol-generating, and scientific evidence supporting that enhanced PPE is necessary to protect OMFs when dealing with suspected and confirmed COVID-19 patients in this current pandemic outbreak. The authors hypothesize that enhanced PPE using respirators (N95 or FFP 2/3) would be sufficient and more effective than standard PPE without respirators or equivalents in protecting dentists and OMF surgeons against COVID-19 during AGPs in suspected and confirmed COVID-19 cases.

## Material and methods

### Protocol and registration

This systematic review followed the Preferred Reporting Items for Systematic Review and Meta-Analysis Protocols(Moher et al., 2015), Figure 1, in combination with the Network Meta-Analyses of Health Care Interventions, and was registered in PROSPERO with No. CRD42020192912 (Essam Ahmed Al-Moraissi, 2020).

**Figure 1:**
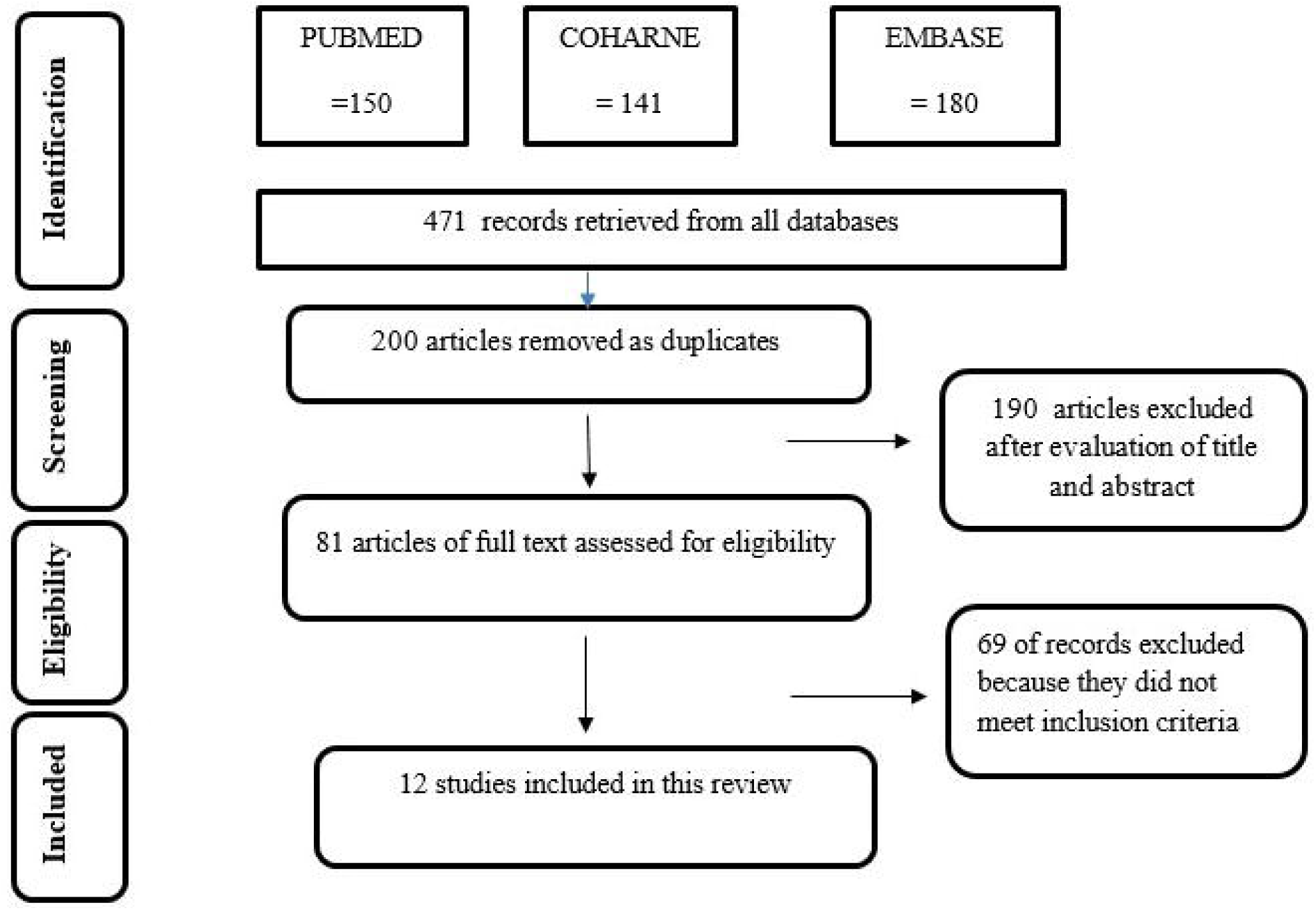
Illustration of PRISMA flow diagram regarding database search strategy.

### Focused questions

1. Do dental and maxillofacial surgical procedures generate bioaerosols (and if so, which ones), which can result in transmission of COVID-19?
2. Are aerosolized airborne droplets (and to which extent is splatter) in dental and maxillofacial surgical procedures infective?
3. Is additional standard personal protective equipment an essential to prevent spreading of COVID-19 during aerosol generating dental and maxillofacial procedures?

### Search strategy

Relevant randomized controlled trials (RCTs), regardless of language and publication date, were retrieved by a systematic search of MEDLINE, EMBASE, CINAHL, the Cochrane Central Registry of Controlled Trials (CENTRAL), and SCOPUS from the inception of each database to the end of May2020 (Additional file 1).

### Selection Criteria

Based on objective of this rapid systematic review, there were 2 inclusion criteria based on the research question as the following :

1. PICOS : Population (P): adult dental health care workers (defined as workers in a health care setting that could be exposed to patients with acute respiratory illness), such as oral and maxillofacial surgeons, dentists or dental assistants who are performing AGPs for unknown, negative, suspected or positive covid-19 patients. Intervention (I): enhanced PPE include respirators such as N95 (certified by the National Institute for Occupational Safety and Health (NIOSH)), FFP2/FFP3 or powered air purifying respirator (PAPR), glove, water proof long sleeved gown, full face shield, head cap and overall cover. Comparator (C): Standard PPE including surgical face mask (certified for use as a medical Device), cloth mask, not wearing any type of facial protections. Outcomes (O): The primary outcome was effectiveness of PPE against covid-19. Study design (S): all literature sources discussing effectiveness of PPE against COVID-19, SARS CoV2, MERS. Additionally, national and international societies’ recommendations, guidelines on using PPE for dental health care workers during the COVID-19 pandemic outbreak were included. Inpatients and outpatients.
2. PICOS: Population (P): patients underwent dental and oral and maxillofacial or orthopedic osteotomies using high speed devices. Intervention (I): high speed devices (C): not applicable. Outcomes (O): detection of aerosols and splatter and count of bacteria Study design (S): all clinical human, cadaver and in vitro studies were be included

### Data extraction

Data extraction was done by 2 reviewers independently. The following information was extracted : authors, type of study, how outcomes were measured, type of surgical procedures, type of surgical instruments used, conclusion, evidence of aerosol generation, evidence of transmission risk and type of microbial species

### Assessment of risk of bias

Because there was extreme heterogeneity among the included studies, risk of bias assessment was not conducted.

### Synthesis of results

Because there was wide heterogeneity among the included studies that investigated the potential aerosolization during dental, OMF and orthopaedic surgeries. Also there was no included study comparing respirators to surgical masks during dental and OMF AGPs for protection against COVID-19 transmission. Thus, results of the current study were presented narratively to answer the prior author clinical questions notably : 1)do dental and maxillofacial surgical procedures generate bioaerosols (and if so, which ones), which can result in transmission of COVID-19?; 2) Are aerosolized airborne droplets (and to which extent is splatter) in dental and maxillofacial surgical procedures infective?; 3) Is additional standard personal protective equipment an essential to prevent spreading of COVID-19 during aerosol generating dental and maxillofacial procedures?

## Results

Based on the literature search, 263 articles were identified overall. Out of these, only 12 studies assessed the potential aerosolization during dental, oral and orthopaedic surgeries were included. Figure 1 is a flow chart on the process of article evaluation for inclusion in the present systematic review.(Earnest et al., 1991; Bentley et al., 1994; Bennett et al., 2000; Nogler et al., 2001a; Nogler et al., 2001b; Nogler et al., 2003; Rautemaa et al., 2006; Ishihama et al., 2008; Ishihama et al., 2009; Yamada et al., 2011; Veena et al., 2015; Singh et al., 2016; Pluim et al., 2018) There was no included study comparing respirators to surgical masks during dental, oral and maxillofacial AGPs for protection against COVID-19 transmission.

### Overall evidence regarding whether dental and maxillofacial surgical procedures generating bioaerosols can result in transmission of COVID-19

There is moderate evidence that ultrasonic scaling and bone drilling using high speed rotary instruments produces respirable aerosols. Additionally, there is very weak/inconclusive evidence to support the creation of infectious aerosols during dental procedures

## Discussion

COVID-19 as a pandemic is currently far from being contained in a majority of countries and represents a serious potential threat to health care workers (HCW), who are disproportionately affected to a higher degree during the current pandemic outbreak (Wang et al., 2020; Zimmermann et al., 2020) Infections rates among HCWs vary greatly depending on literature, with ranges between 1.6 – 3.8 in countries which succeeded to contain the first wave of the pandemic, such as e.g. Germany or China (Korth et al., 2020; Schmidt et al., 2020; Wang et al., 2020), which stands in conspicuous contrast to countries with outbreaks getting out of control such as Italy (Remuzzi et al., 2020). Especially in the context of AGPs, Covid-19 has also reawakened the longstanding debate about the extent to which common respiratory viruses, now including SARS-CoV-2, are transmitted via respiratory droplets vs aerosols, so called airborne transmission(Klompas et al., 2020). The role of human exhaled droplets, direct contact and fomites (the latter ones being regarded as a less relevant way of transmission) for the transmission(Sommerstein et al., 2020) of SARS-CoV2 and COVID-19, respectively, has been primarily addressed by standard and adapted hygienic measures (e.g. patient screening by point of care assessment, individual risk assessment and rapid testing, distancing, hand hygiene, environmental surface disinfection etc.) combined with standard and/or enhanced PPE(Bartoszko et al., 2020; Zimmermann et al., 2020) to protect HCW when dealing with suspected and confirmed COVID-19 patients. According to available epidemiological data, SARS-CoV2 has a higher transmissibility than SARS-CoV or MERS-CoV (Chen, 2020) modification of standard precaution and infection control regimen targeted towards SARS-CoV2(Ge et al., 2020), therefore, may be crucial.

### Do dental and maxillofacial surgical procedures generate bioaerosols (and if so, which ones), which can result in transmission of COVID-19?

As, regarding focused question 1 *(“do dental and maxillofacial surgical procedures generate bioaerosols - and if so, which ones-, which can result in transmission of COVID-19?”)*, especially potential airborne transmission of COVID-19 via aerosols (Wax et al., 2020), as fourth way of transmission(Sommerstein et al., 2020), though controversially discussed, is considered as a significant risk (Schwartz et al., 2020), particularly for all those medical professions working in close vicinity to the bronchotracheal, nasal and paranasal, oral and oropharyngeal system(Patel et al., 2020; Zimmermann et al., 2020). Although conclusive data regarding concrete numbers of incidence among dental and OMFS HCWs are basically lacking, there are some reports indicating that dentists and OMFS are among those at elevated risk(Ge et al., 2020; Warnakulasuriya, 2020) for transmission by patients including asymptomatic or prior to onset patients. According to Meng et al., 2020, the infection rate among dental 169 Chinese dental staff, in Wuhan city who treated more than 700 emergencies using medical masks for personal protection was as high as 5.3 % (Meng et al., 2020), which is at least above the rate of 2.7% for HCWs tested seropositive for SARS-CoV2 in a German tertiary center in a pandemic hotspot as described above(Korth et al., 2020; Schmidt et al., 2020) SARS-CoV2 has an even higher transmissibility than SARS-CoV or MERS-CoV(Chen, 2020) and both SARS-1 and MERS coronaviruses have shown high transmissibility, both with proportions of infected ranging overall from 13 to 43%, based on country-specific data. For individual outbreaks, up to 59% of affected individuals were HCWs(Sommerstein et al., 2020). Therefore, it rather likely that the number of infections among oral surgeons and OMFS will rise either with the progress of the pandemic and/or in the wake of reverting to normal business. In addition, considering potential aerosol transmission, due to the specific characteristics of their working environment, oral surgeons and OMFS, may contribute themselves inadvertently to cross-transmit COVID-19 from patient to patient and HCWs by aerosol transmission during and for some time after performing AGPs, especially when there is an exposure to high concentrations of aerosols in a relatively closed environment such as in surgeries(Ge et al., 2020; Morawska et al., 2020) Though dentists and oral surgeons have been recommended to avoid or minimize operations that can produce droplets or aerosols (Meng et al., 2020)under recent lockdown conditions, by now many countries have reverted to medical business as usual, thus many HCWs by now are facing a second wave of the pandemic or risk further hampering of comprehensive patient treatment(European Centre for Disease Prevention and Control, 2020; Zimmermann et al., 2020). For dentists, OMFS, ENT etc. working closely to the oropharyngeal and tracheobronchial system, determining whether droplets or aerosols predominate in the transmission of SARS-CoV-2, therefore, turns out to be a crucial question with high implications. If – as it is considered highly probable by the proponents of airborne transmission(Morawska et al., 2020) - SARSCoV-2 is in fact transmitted via aerosols (which can remain suspended in the air for prolonged periods such as pollens(Klompas et al., 2020),medical masks would be inadequate (Morawska et al., 2020)because aerosols can both penetrate and circumnavigate masks. Face shields, too would provide only partial protection as they leave open gaps between the shield and the HCW, and 6 feet of separation would not provide protection from aerosols that remain suspended in the air or are carried by currents (Klompas et al., 2020; Morawska et al., 2020). As a consequence, understanding aerosol transmission and its implications in dentistry are essential(Klompas et al., 2020), as oral surgery environments with AGPs convey high risk of aerosolised transmission(Zemouri et al., 2017; Zimmermann et al., 2020). Zemouri et al. with high-speed drilling, water-air syringe, ultrasonic scaling and piezosurgery generally considered to be high risk transmitters(Ge et al., 2020; Warnakulasuriya, 2020; Zimmermann et al., 2020). In OMFS, tracheostomy, tracheostomy care, airway suctioning, abscess drainage and wound irrigation (e.g. hydro-jet lavage) need to be added to this list according to the WHO recommendations(2020, 2020) based on prior experiences with SARS-CoV-1(Sommerstein et al., 2020). Although the production of aerosols during these AGPS goes generally accepted, there is overall only weak to moderate evidence that these aerosols his will in fact cause aerosol-based transmission. Whereas a majority of experimental data usually based on bacterial tests e.g. during use of dental high speed rotary instruments and/or ultrasonic scalers (Table 1) consider evidence of transmission risk during AGPs either as basically unclear(Earnest et al., 1991; Ishihama et al., 2009) (Bennett et al., 2000; Rautemaa et al., 2006). E.g., Ishihama et al 2008 assessed high speed rotary instruments during surgery of impacted third molars and found only indirect evidence supporting generating aerosols during oral surgery (Ishihama et al., 2009)

**Table 1:**
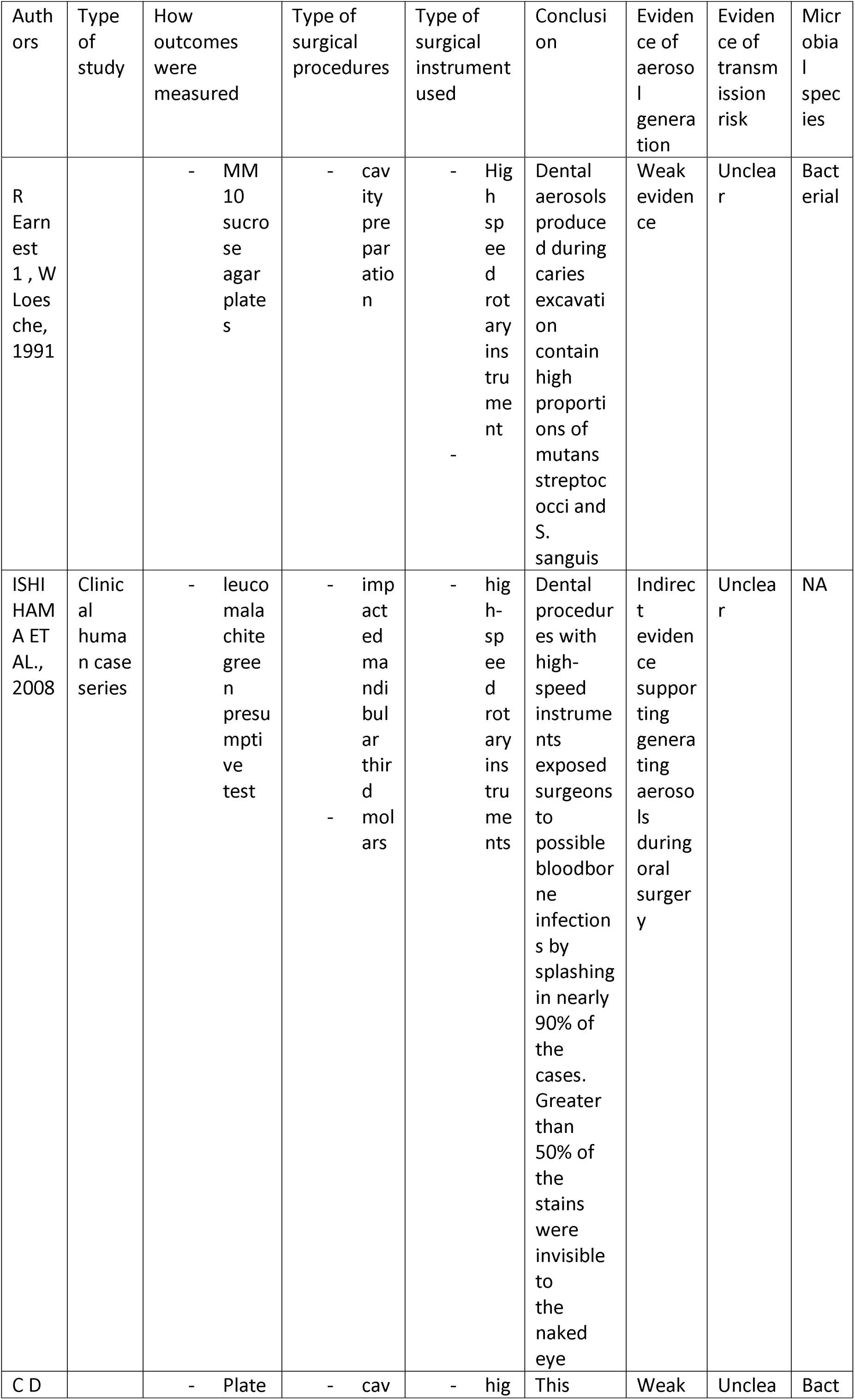
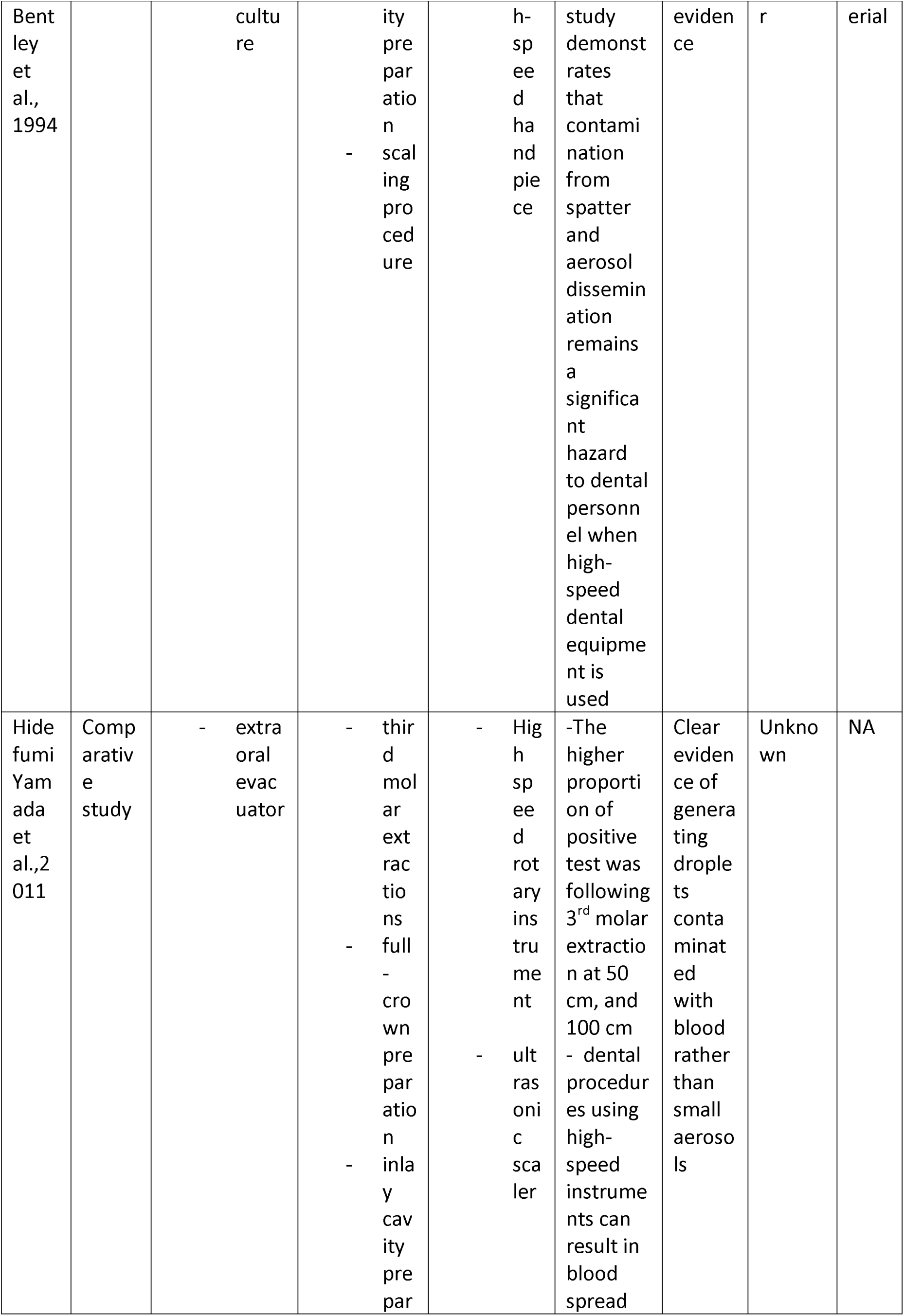
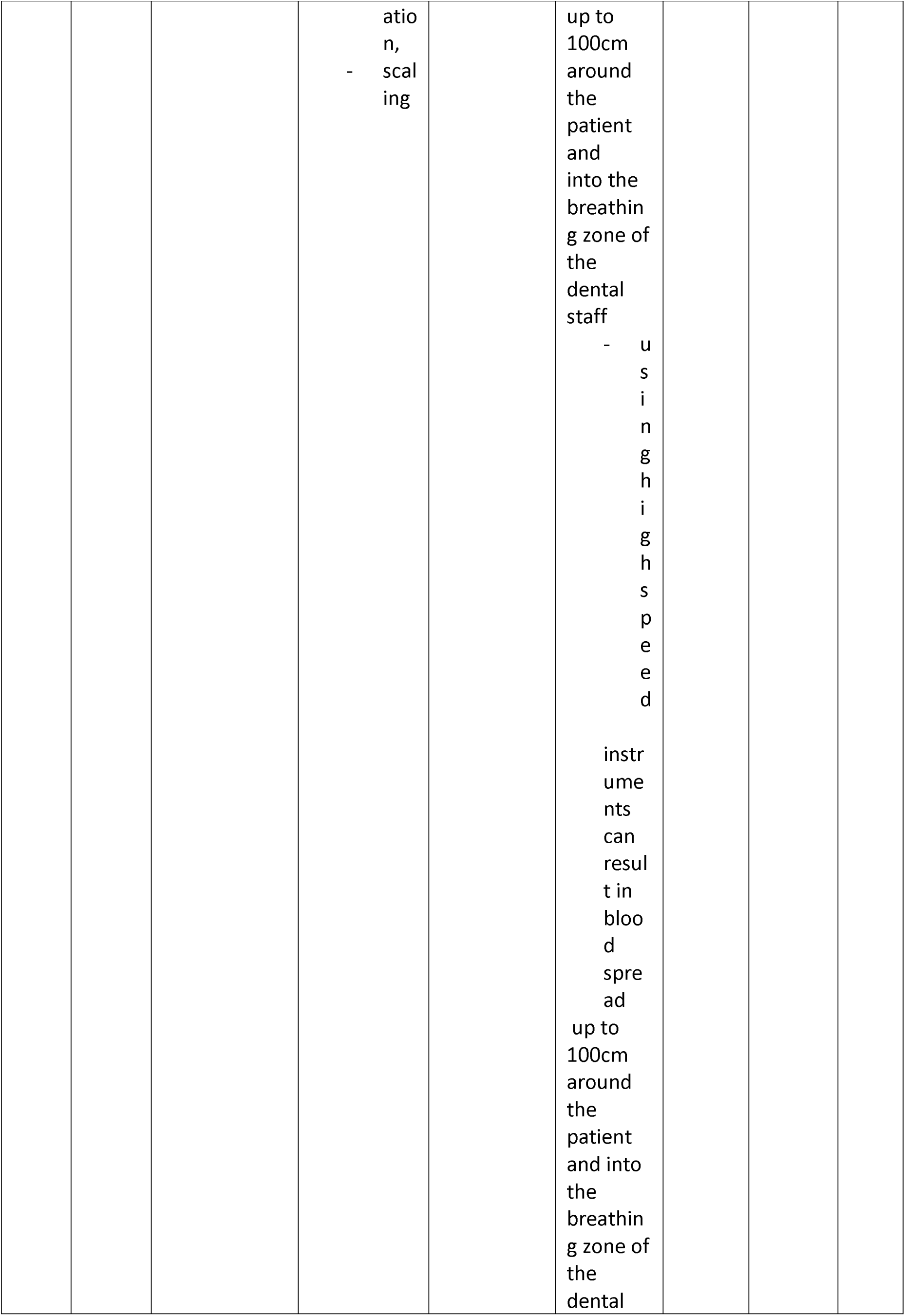
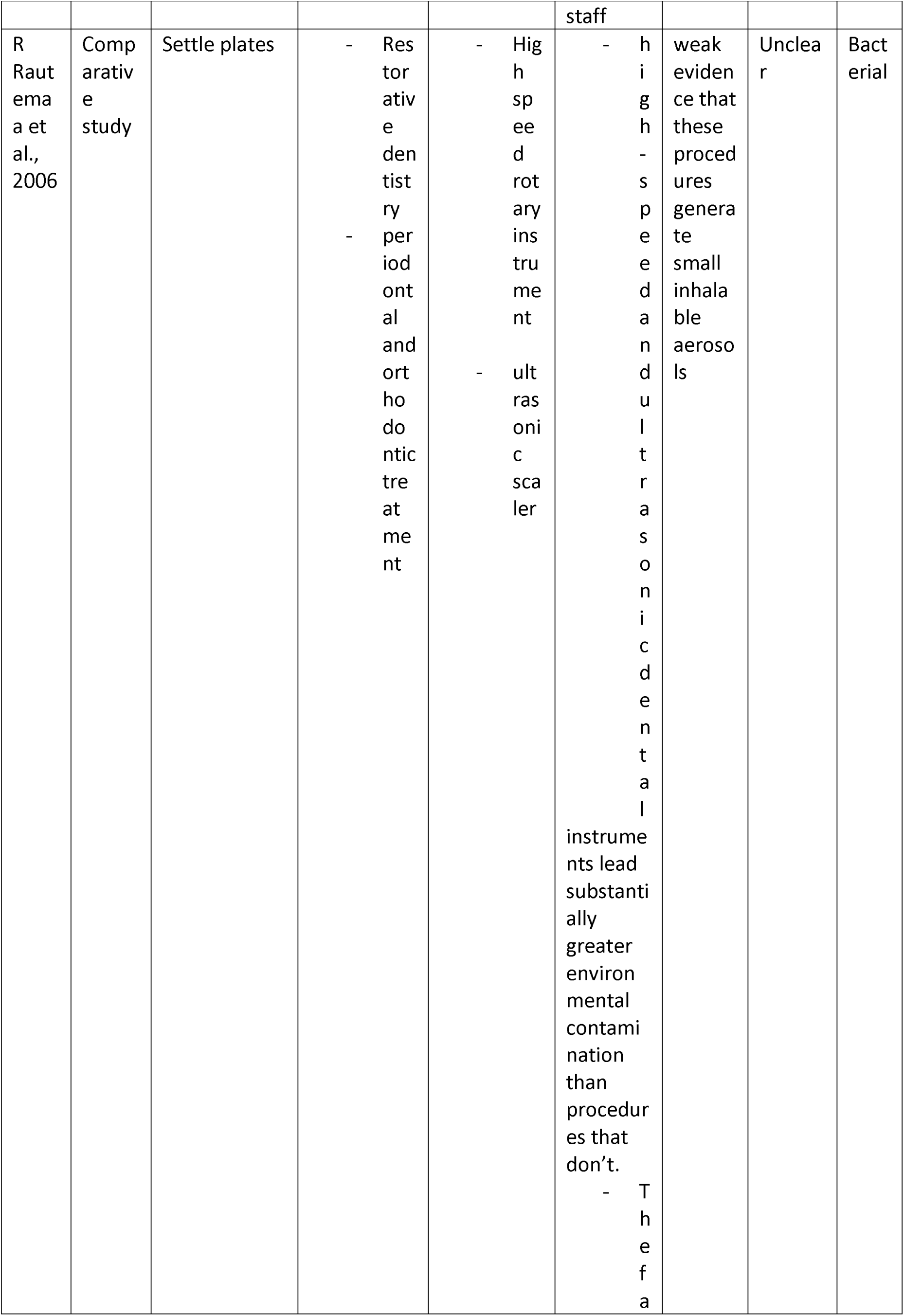
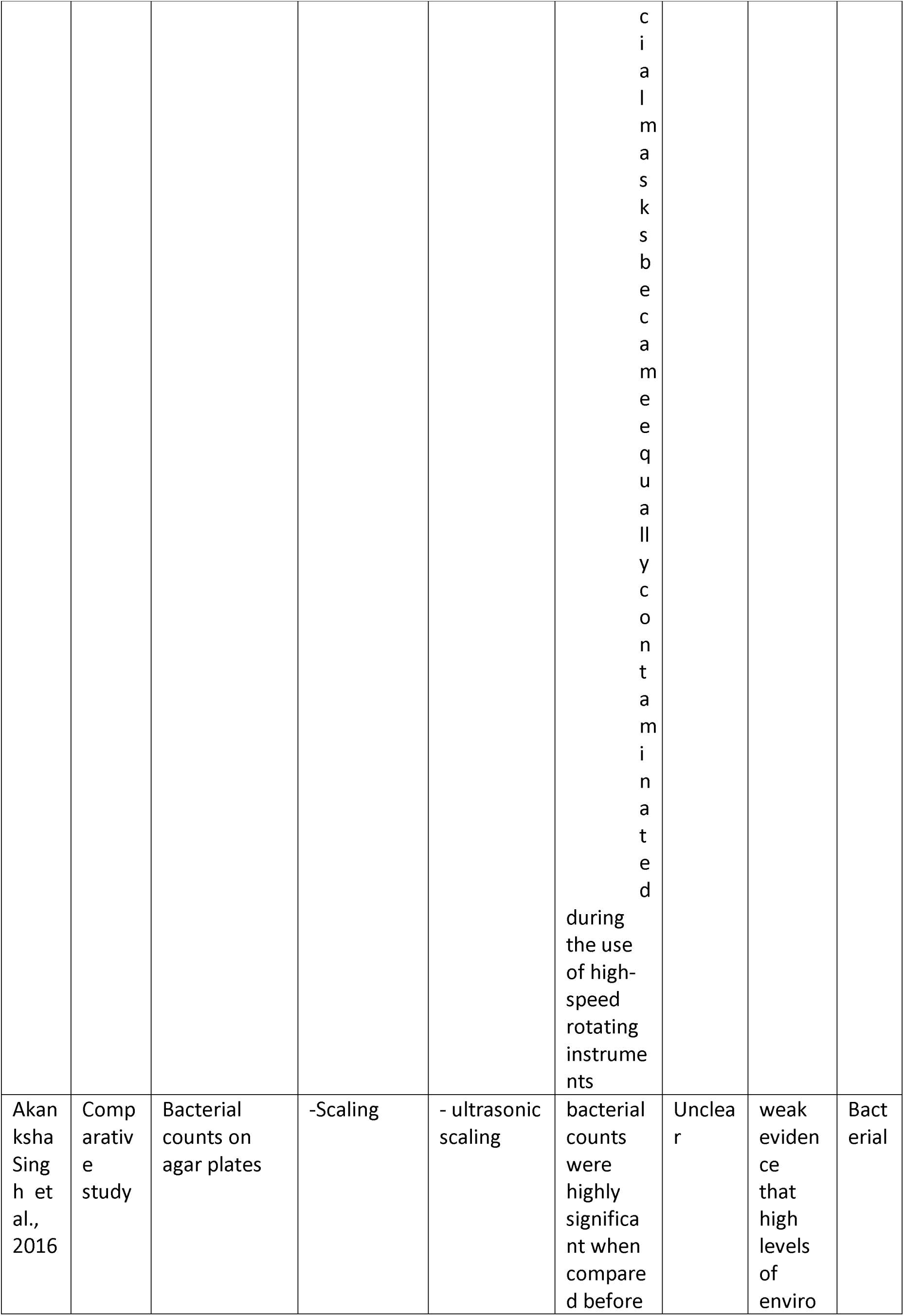
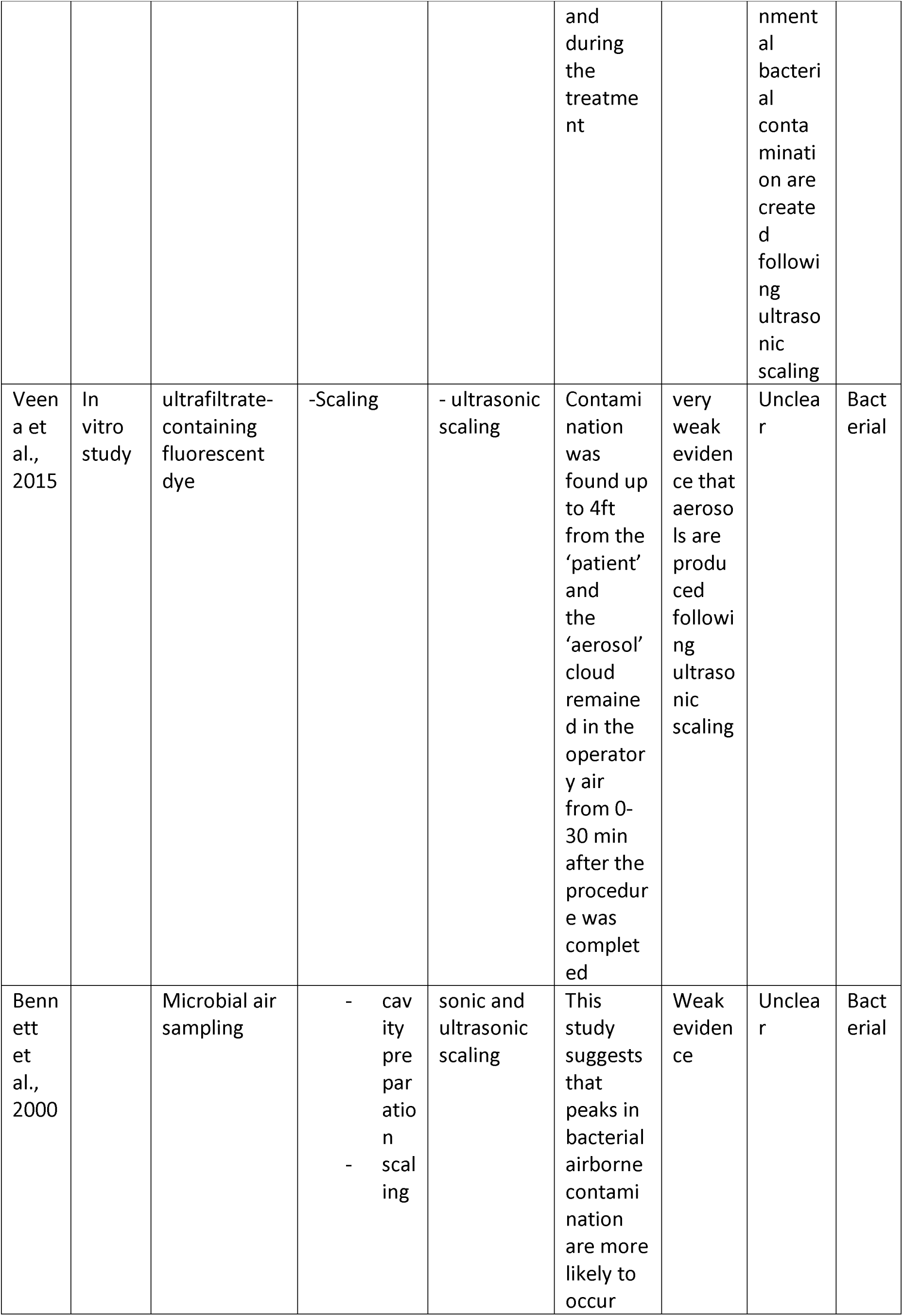
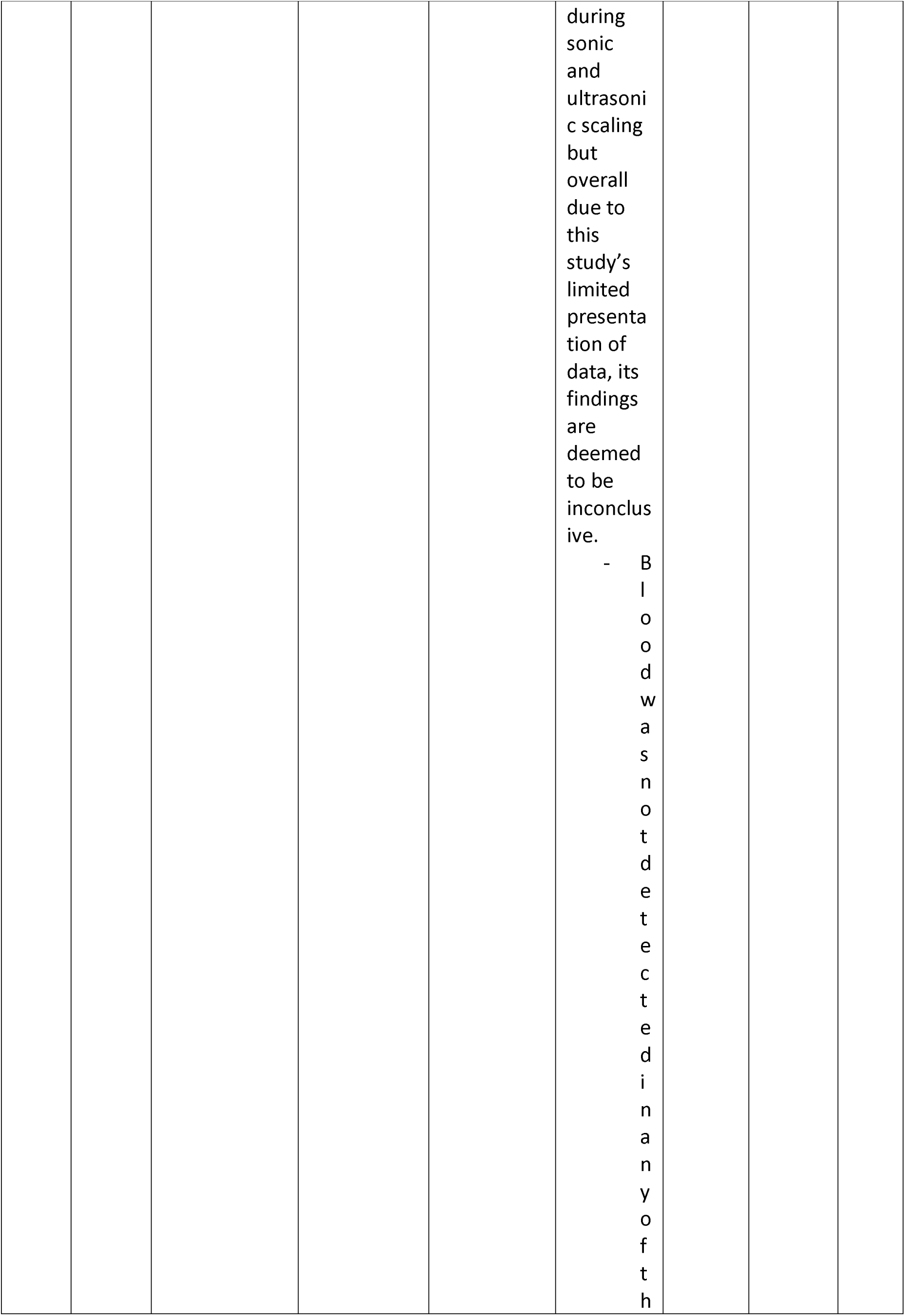
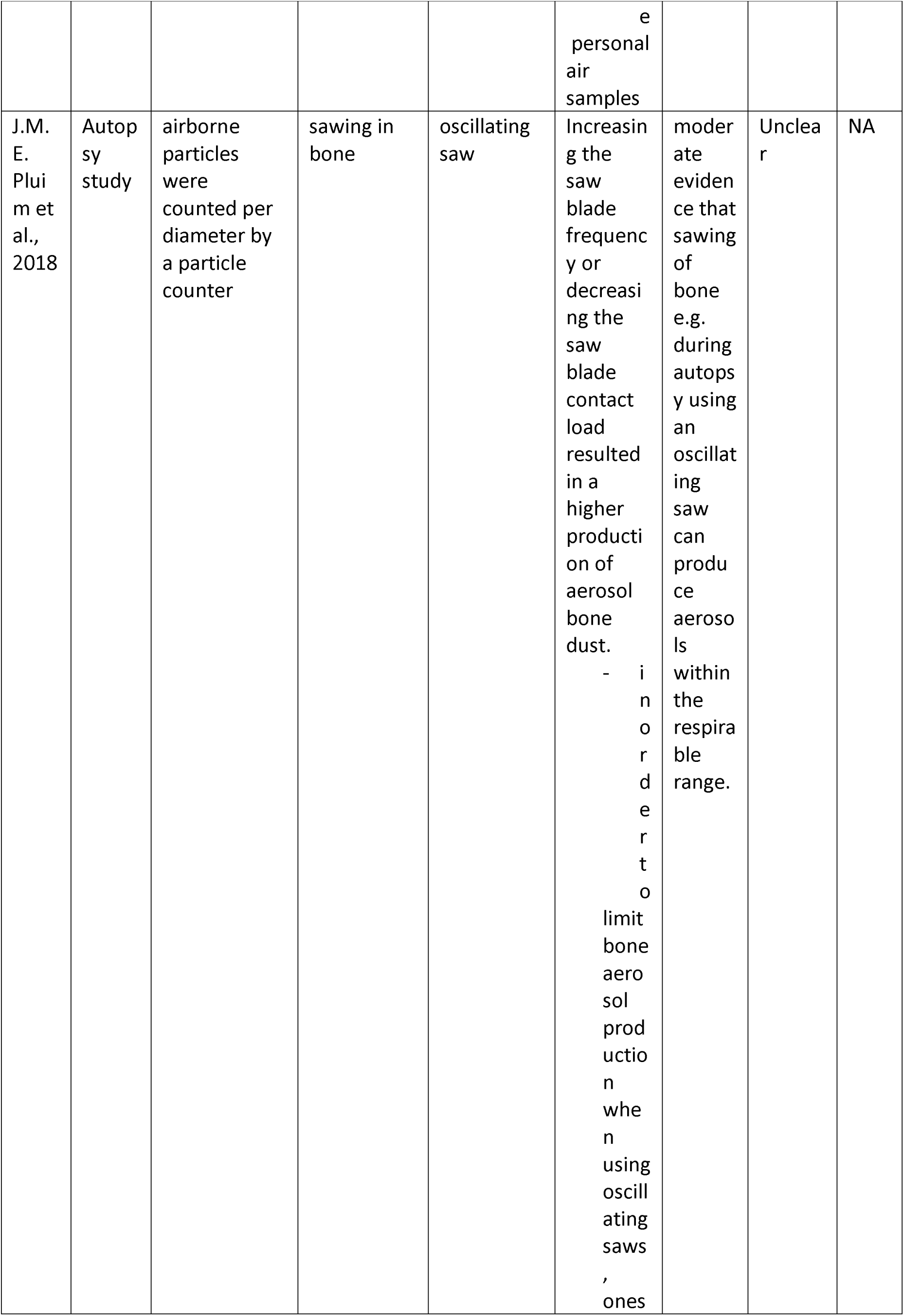
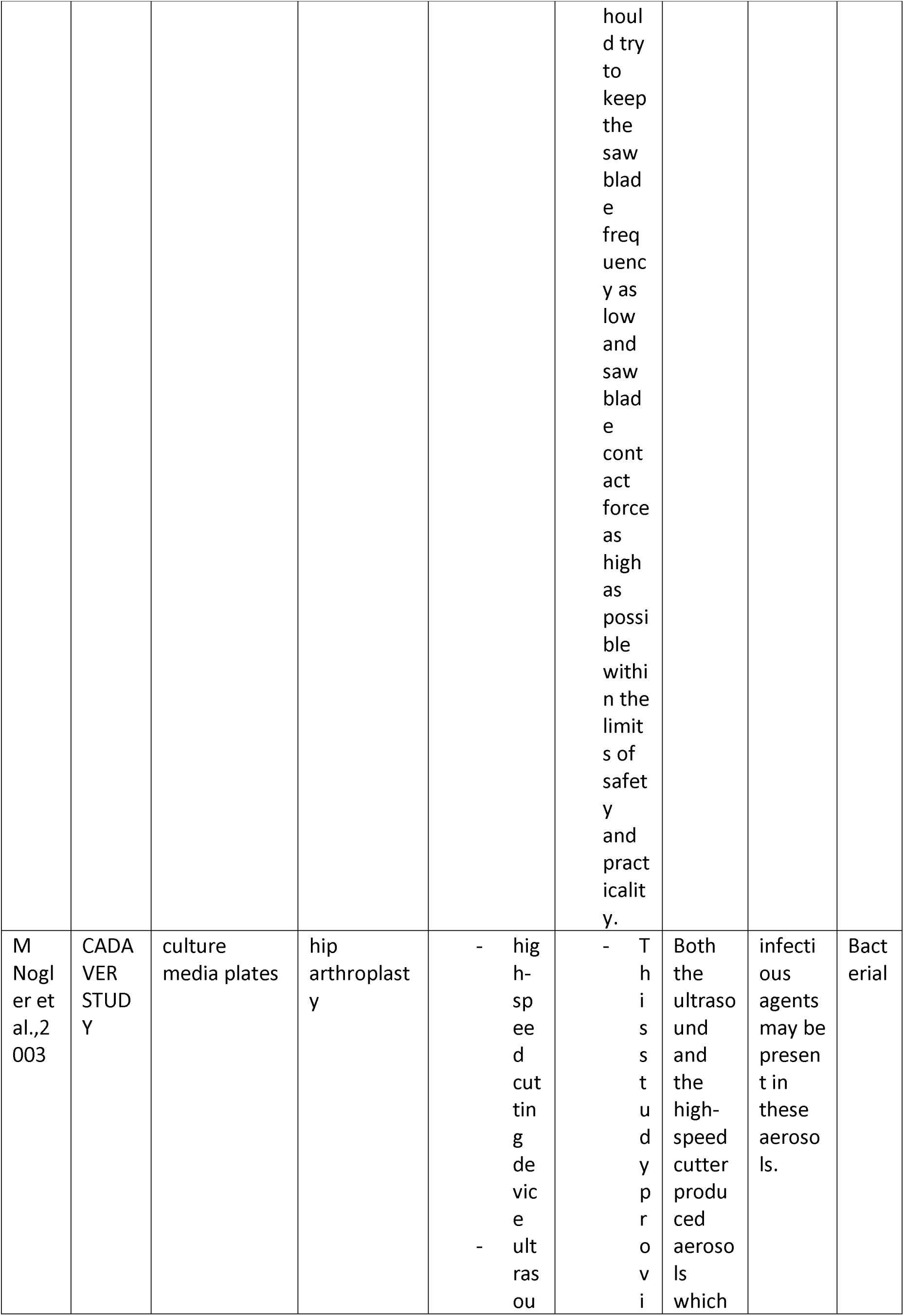
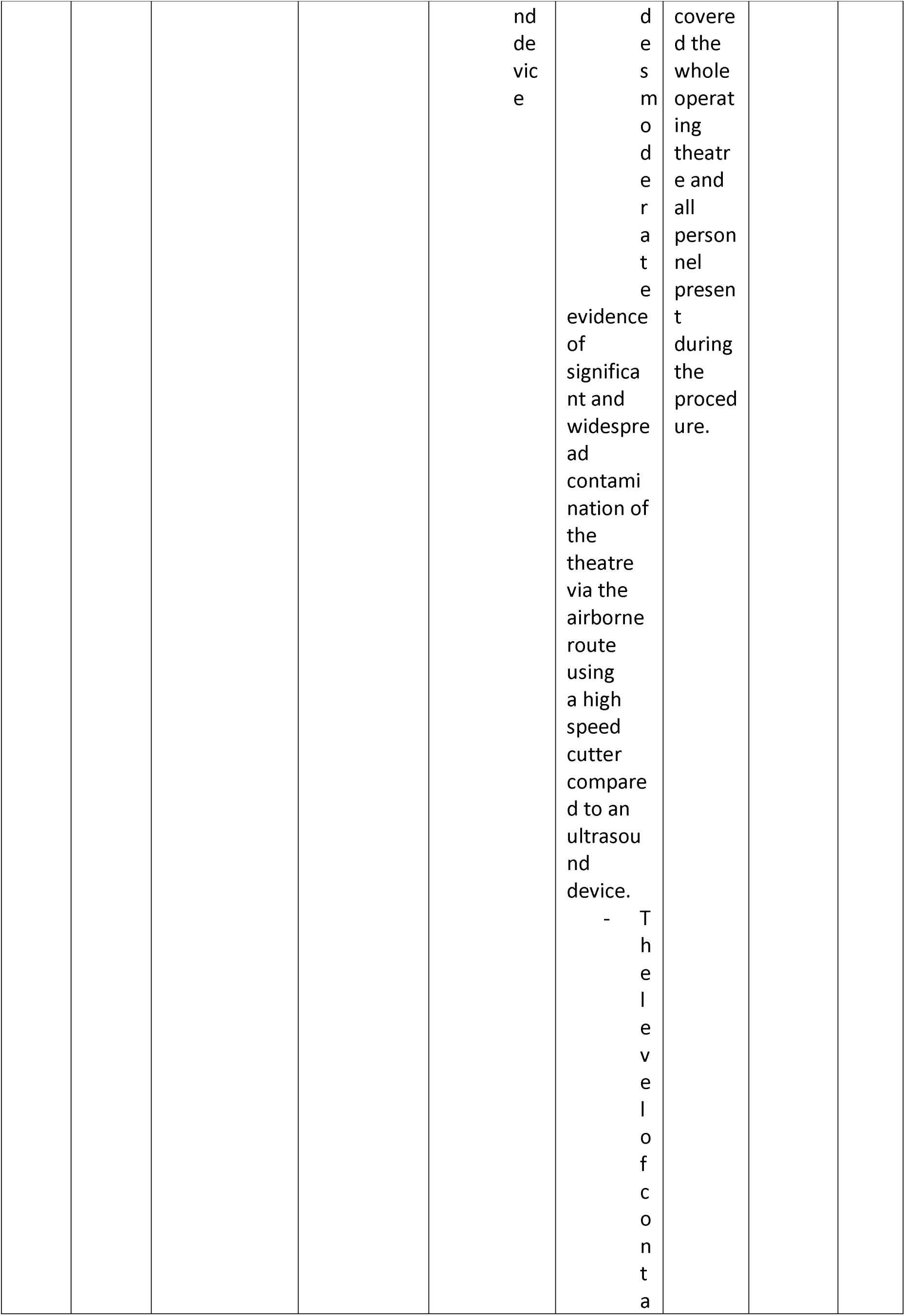
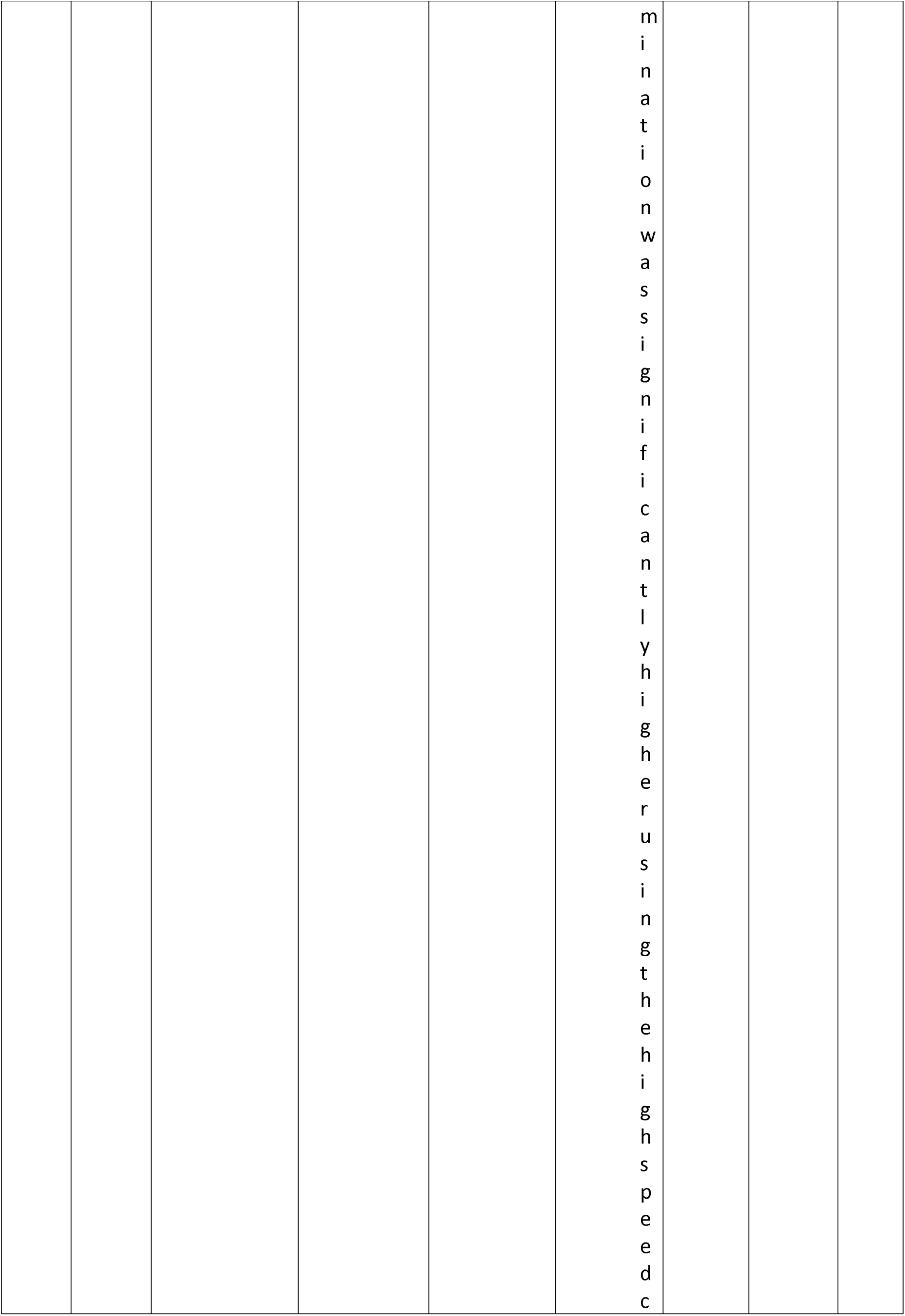
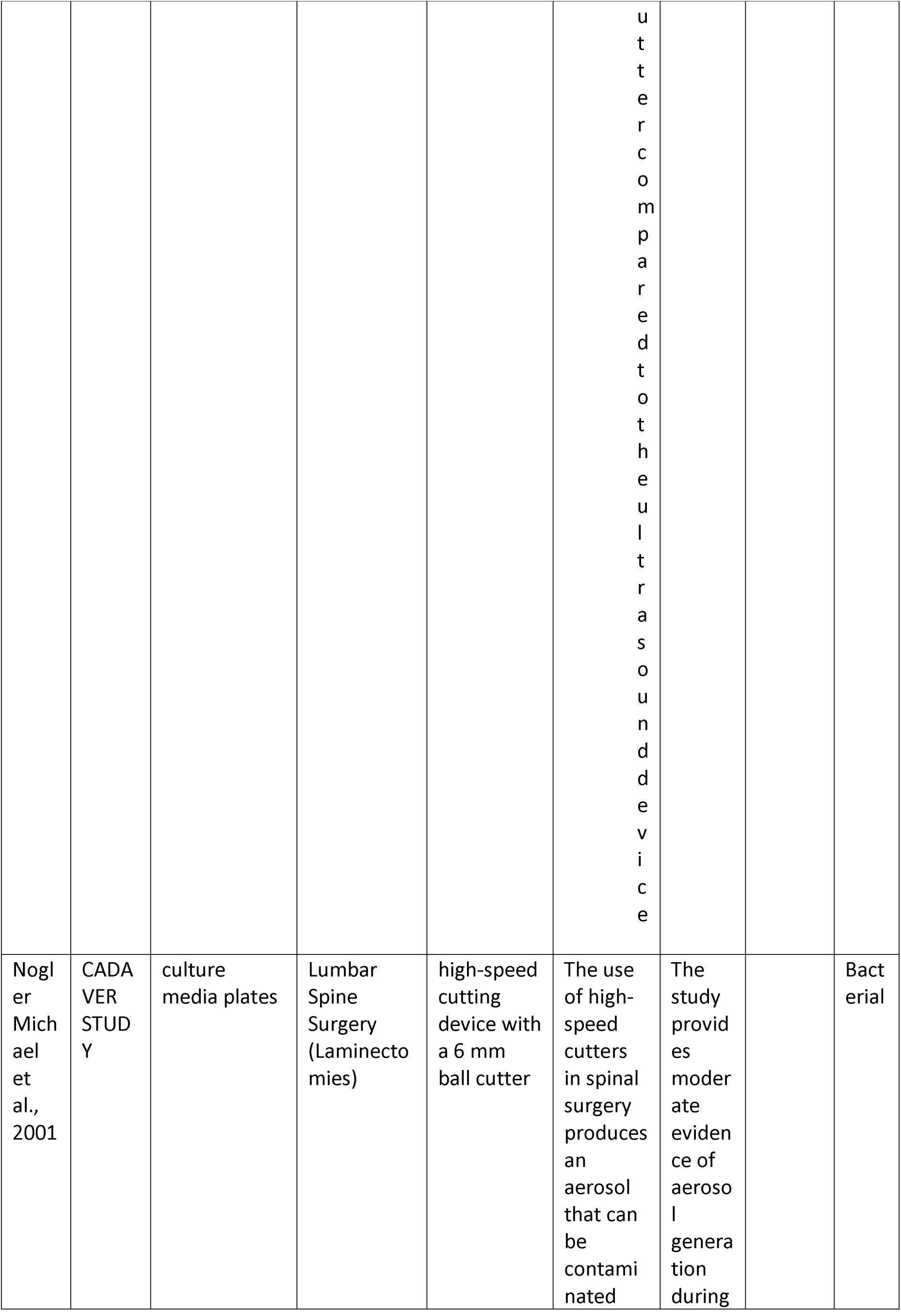
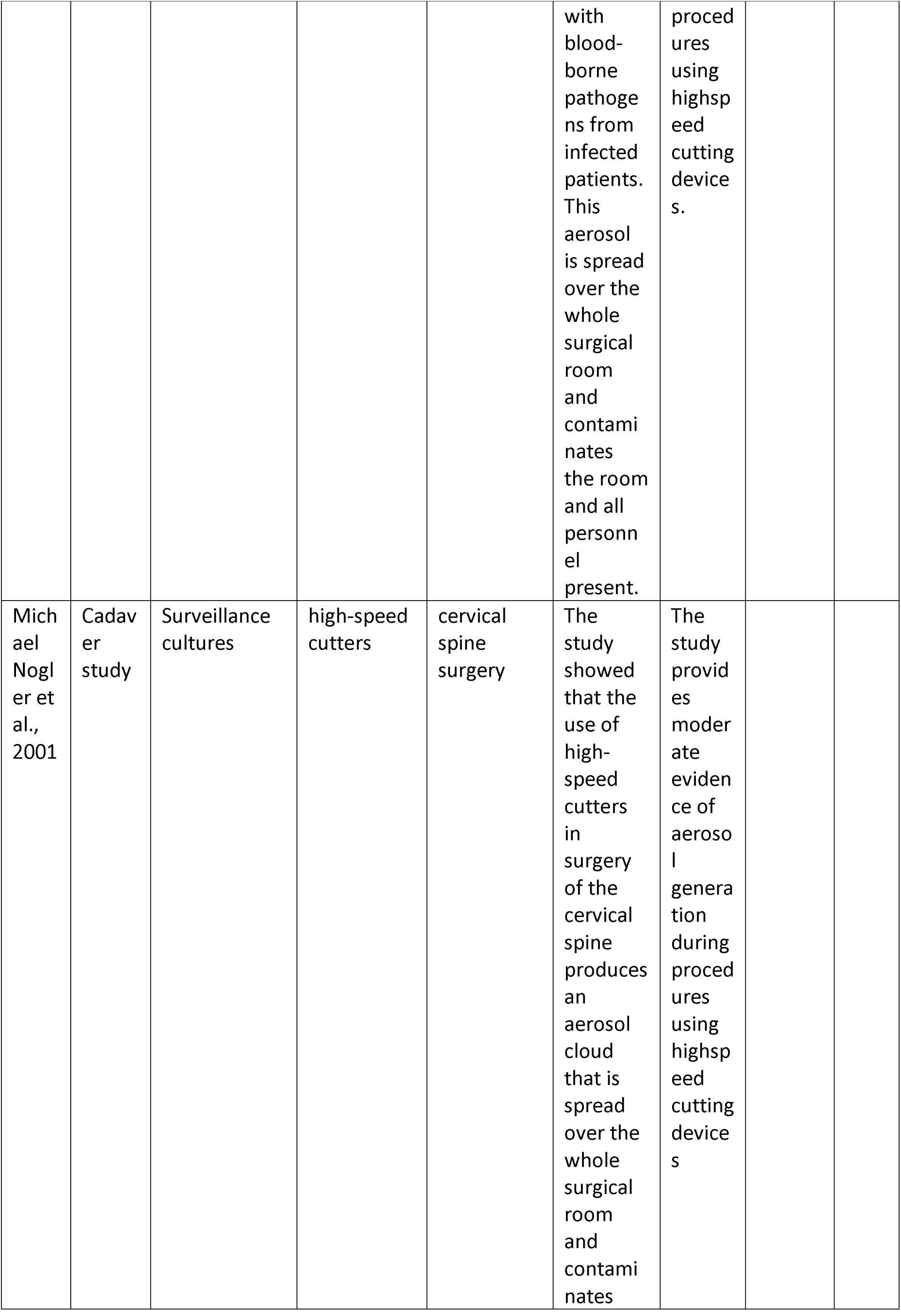
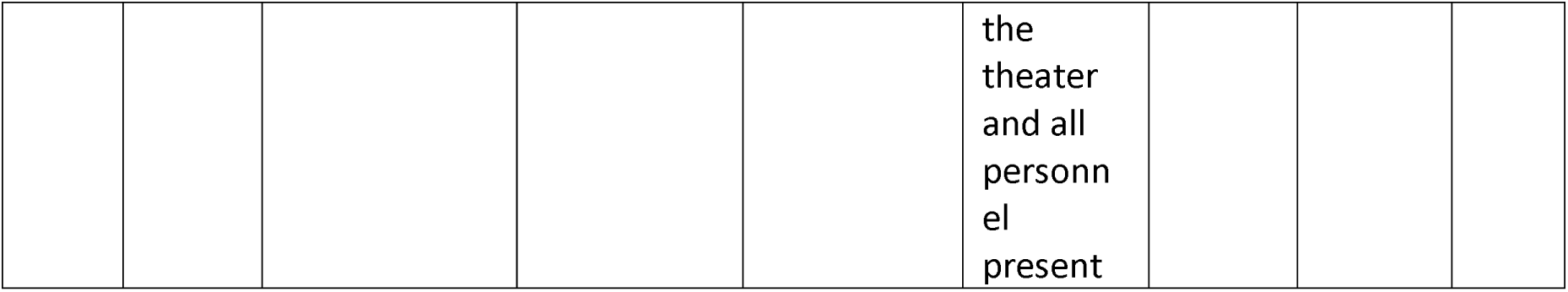
characteristics of included studies

Hidefumi Yamada et al. 2011, evaluating both high speed rotary instruments and ultrasonic scalers concluded that there is clear evidence of generating droplets contaminated with blood rather than small aerosols (Yamada et al., 2011). Regarding extraoral osteotomies, Nogler et al. 2001 assessed a highspeed cutter in spinal surgery, which produced an aerosol covering the complete operating theatre and all personnel, which can be contaminated with blood-borne pathogens from infected patients(Nogler et al., 2001a). Based on bacterial material, this study provided moderate evidence of aerosol generation (Nogler et al., 2001a)

In 2003 these authors compared ultrasound and highspeed bone cutting. Both the ultrasound and the highspeed cutter produced aerosols which covered the operating theatre and all personnel present. The highspeed cutter caused significantly higher contamination compared to the ultrasound device (Nogler et al., 2001a). These findings are in line with JME Pluim et al. 2018, who found moderate evidence that sawing of bone e.g. during autopsy when using an oscillating saw can produce aerosols within the respirable range (Pluim et al., 2018). Therefore, aerosol formation during OMFS bone cutting procedures needs to be considered as a potential risk factor and the question arises whether there are potential infectious agents present in these aerosols.

### Are aerosolized airborne droplets (and to which extent is splatter) in dental and maxillofacial surgical procedures infective?

In this context, regarding focused question 2 *(“Are aerosolized airborne droplets - and to which extent is splatter - in dental and maxillofacial surgical procedures infective?”)* it needs to be stressed that already normal breathing and coughing will produce aerosols (i.e., even without AGPs as postulated by the proponents of airborne transmission (Morawska et al., 2020). Klompas et al, however, pointed out that this does not mean aerosols his will in fact cause aerosol-based transmission (Klompas et al., 2020) as this depends – besides route of exposure - on factors such as the size of inoculum, duration of exposure and host defenses(Klompas et al., 2020). Low reproduction numbers of COVID-19 (rather similar to influenza, i.e. R_0_ ≈ 2 as opposed to classical airborne viruses such as measles, with R_0_ ≈ 18) (Sommerstein et al., 2020)indicate that either a high virus-load is required, or aerosols are not the dominant mode of transmission (Klompas et al., 2020). Although some possible superspreading events been reported for both SARS-CoV-1 and now SARS-Cov-2, such as 53/61 attendees of a choir infected by one singer(Frieden et al., 2020) (Hamner L, 2020), but, this does not necessarily support aerosol transmission (Frieden et al., 2020), but may be explained by infection via droplets and contact alternatively, as well(Sommerstein et al., 2020). At any rate, as Morawska & Milton state in their recent commentary supported by 239 scientists, the precautionary principle should apply and a heightened awareness for airborne transmission is required(Morawska et al., 2020). During the SARS-1 epidemic, higher infection rates have been well documented for manual ventilation before and during intubation, tracheotomy and non-invasive ventilation(Tran et al., 2013; Sommerstein et al., 2020). In analogy, close vicinity and prolonged exposure during dental and OMFS AGPs, in combination with poor ventilation, therefore, may play a relevant role in transmission of SARS-CoV-2. According to Cummings et al., based on the cluster randomized Respiratory Protection Effectiveness Clinical Trial (ResPECT) in 4689 health care personal seasons assessed between 2011 and 2016, the risk to become infected with an endemic coronavirus was increased approximatively twofold with exposures to AGP (al, 2020). Though SARS-CoV-2 may behave differently, this indicates a potential and at least clinically relevant airborne route of transmission. At present, from an evidence based point of view, current understanding is considered to be still limited and there is a lack of higher level experimental data proving or disproving droplet vs aerosol-based transmission of SARS-CoV-2 particularly in well-ventilated spaces(Klompas et al., 2020). Though based on weak evidence, however, additional control measures against airbone transmission - at least during AGPS - are reasonable to reduce risk of infection for dental and OMFS HCWs. As far as oral examination and oral surgeries are concerned, such as extractions, incisions and drainage of dental abscesses, spread via droplets and splatters is certainly prevalent in contrast to aerosolization(Yamada et al., 2011). Low speed bone drilling or sawing under saline solution cooling, however, will inevitably produce droplets, which may spread further than 2□m with strong directional airflow support(Sommerstein et al., 2020). Basically, these droplets then may even turn into aerosols by evaporating when suspended long enough in the air, i.e., turning into microdroplets < 5μm faster than they settle, thus forming true “droplet nuclei” or aerosols (Sommerstein et al., 2020). Basically, particles ≤10□µm are considered respirable particles which are capable of reaching the lower airways, whereas particles with 10– 100□µm are considered inspirable particles, i.e., limited to reach the upper airways (Sommerstein et al., 2020).As viral RNA (though no viable virus) has been detected in the air associated with droplets smaller than, the droplets may maintain infectivity(Morawska et al., 2020). SARS-CoV has been reported to travel more than six feet(Kutter et al., 2018). There is high probability termed “beyond reasonable doubt”(Morawska et al., 2020), that e.g. patients’ breathing, talking and less likely coughing (Sommerstein et al., 2020)e.g. during surgery may cause a mix of potentially infective droplets and aerosols. Microdroplets small enough to remain aloft in air thus pose a risk of exposure at distances beyond 1 to 2 m from an infected patient(Morawska et al., 2020), and aerosols are estimated to travel between up to 4.5 m (Loh et al., 2020) and 27 feet (around 8 m), or room scale (Sommerstein et al., 2020), respectively, and stay viable for hours (Liu et al., 2020). In this context, factors such as indoor air velocities need to be considered, too, which may allow a 5 μm droplet to travel tens of meters, explaining e.g. some spatial patterns of the SARS-CoV-1 epidemic with high likelihood (Morawska et al., 2020), though alternative non-airborne ways of spreading need to be considered, equally (Klompas et al., 2020). however, this does not prove COVID-19 to be a truly aerosol-transmitted disease. In contrast to so far non-proven airborne transmission, transmittable virus had been cultured from fomite samples in a MERS-CoV outbreak in South Korea (Bin et al., 2016) resulting in guidelines unanimously requiring contact isolation in addition to droplet precautions and propagating strict hand hygiene (Sommerstein et al., 2020)

In contrast to these considerations mentioned above by the proponents of airborne transmission, the World Health organization (2020) and most public health organizations do not recognize airborne transmission except for AGPs (Morawska et al., 2020). Considering low R0-rates and the striking success of social distancing(Sommerstein et al., 2020), the focus usually lies instead on prevention of contact contamination and direct droplet transmission, which itself poses a special threat to dental and OMFS HCWs. SARS-CoV2 has been found in infected saliva(To et al., 2020)

In addition, a high expression of angiotensin-converting enzyme II (ACE2), representing the cell receptor for SARS-CoV2 infection, has been reported in the oral cavity, especially in the epithelial cells of the tongue (Xu et al., 2020), turning the oral cavity into a potential high risk transmitter (Ge et al., 2020), even in case airborne transmission should tun out to be neglectable. This local virus load also explains the potential of preprocedural mouth rinsing (e.g. chlorhexidine CHX), which leads to a mean reduction of 68.4% colony forming units in dental aerosols (Marui et al., 2019) and which is proven to be efficient against several infectious viruses (Ge et al., 2020). Efficacy may be augmented by adding 1% hydrogen peroxide or 0.2% povidone iodine, as SARS-CoV-2 seems to be sensitive to oxidation(Peng et al., 2020; Zimmermann et al., 2020). Nevertheless it should always be kept in mind that airborne transmission via aerosols remains am imponderable threat especially to oral and OMFS surgeons even though so far un-proven (Sommerstein et al., 2020)and still speculative, which may be due to the fact that it is difficult to detect contaminated air, because infectious aerosols are usually extremely dilute, and that it is hard to collect and culture fine particles (Roy et al., 2004). Chad et al. therefore suggested to classify the aerosol transmission of diseases as obligate, preferential, or opportunistic, on the basis of the agent’s capacity to be transmitted and to induce disease through fine-particle aerosols and other routes (Roy et al., 2004; Shiu et al., 2019). Even though orthodox ways of transmission are more likely to occur, “unorthodox” transmission ways may, nevertheless, may gain importance under special conditions, such as AGPs in oral and OMFS surgery.

### Is additional standard personal protective equipment an essential to prevent spreading of COVID-19 during aerosol generating dental and maxillofacial procedures?

As a basic principle, the same precautions should apply for all patients regardless of case status (positive, carrier or negative) during the period of sustained COVID-19 transmission including standard local disinfection and decontamination protocols plus pandemia adapted distancing procedures etc. to effectively limit the concentration of SARS-CoV-2-RNA in aerosols(Li et al., 2004). Though the respective recommendations issued e.g. by the Chief dental officer, dental chambers or respective governmental and health service institutions of the different countries need to be followed by HCWs, many recommendations are heterogeneous and epidemiological data relative to their effectiveness against COVID-19 are limited (Bartoszko et al., 2020). Therefore, from a clinical point of view, the most contingent question arises as to which is an adequate/appropriate PPE for dental and OMFS AGPs and whether this question can be answered from an evidence-based point of view. As a general principle, PPE should be chosen depending on the planned procedure and the infection status of the patient in order to save resources (Zimmermann et al., 2020).So far, according to general consensus, power air-purifying respirators (PAPR), which were scarcely available during the outbreak, so far have not been considered mandatory to safely avoid aerosol-borne transmission in OMFS (Zimmermann et al., 2020).At present, N95/FFP2 for AGPs and N99/FFP3 masks with (Zimmermann et al., 2020)for surgery in infected patients, respectively, are most frequently recommended, instead (BAOMS, 2020; Centers for Disease Control and Prevention, 2020b; European Centre for Disease Prevention and Control, 2020; French Society of Stomatology et al., 2020b; Occupational safety and health administrations, 2020; The American Society of Dentist Anesthesiologists, 2020)

In Spain, which was severely hit by COVID-19, the OMFS society (SECOM-CYC) recommends use of FFP3 and sealed glasses or alternatively face shields for the OR in combination with waterproof surgical gown and gloves plus double disposable surgical cap during AGPs in symptomatic and COVID-19 positive patients (Monje Gil et al., 2020)

Chu et al. (Chu et al., 2020)concluded in their recent systematic review and meta-analysis regarding spread of viruses via aerosols, that respirators would be more protective than medical masks alone. This metanalysis, however, as Klopas et al pointed out, was not based on a direct comparison of N95 respirators vs. medical masks and mostly included studies on SARS-CoV1 and MERS virus. In addition, it was based on a post-hoc bayesisan analysis of two independent analyses, one, however, on N95 respirators vs no masks and the other on medical masks vs no masks (Klompas et al., 2020).In contrast, Bartoszko et al. in their recently published systematic review and meta-analysis, regarding use of medical masks vs N95 respirators in preventing laboratory confirmed viral infection and respiratory illness specifically in HCWs, analyzed four RCTs including coronavirus and concluded that the use of medial masks did not increase the rate of laboratory confirmed respiratory infection (OR1.06) or clinically respiratory illness (OR 1.49). One trial evaluating coronaviruses (though not SARS-CoV-2) found no difference between the two groups (p=0.49). This may not only apply for routine care and non AGPs, but according to a case report comprising 35/41 HCWs (Ng K et al. 2002, Ann Intern Med) also for AGPs. All 35 of these HCWs wearing just medical masks when exposed to SARS-CoV-2 during high risk AGPs were tested seronegative after 14 days. It therefore may be supposed that high quality standard surgical masks (type II/IIR according to European Norm EN 14683) appear to be comparably effective as FFP2 masks in preventing droplet-associated viral infections of HCWs as reported e.g. from influenza or SARS-CoV-2 (Sommerstein et al., 2020). In this context, is also noteworthy that e.g. in the UK HCWs with and without direct patient contact showed similar incidence rates of COVID-19, implying that community-acquired disease or transmission among co-workers were more likely than nosocomial transmission from infected patients (Hamner L, 2020; Sommerstein et al., 2020) Nevertheless, at least for AGPs, N95 respirators/FFP-2 masks at present are unanimously recommended by national and international guidelines. The underlying rationale most probably relates to high level of viral exposure from droplet clouds rather than transmission by the airborne route (Sommerstein et al., 2020), but is also due to the conspicuous lack of understanding of the detailed mechanisms of SARS-Cov-2 transmission, which may also explain the discrepancy of the recommendation to protect the HCWs with surgical masks versus respirators (Sommerstein et al., 2020). Accordingly, there is inconsistency in recommendations for routine care and non AGPs of COVID-19 (Morawska et al., 2020)as the World Health Organization, Public Health England and the Public Health Agency of Canada recommend the use of medical/surgical masks for non-AGPS (Public Health Agency of Canada, 2020; Public Health England, 2020a; World Health Organization, 2020) in contrast to several societies and national associations (such as the U.S Centers for Disease Control and Prevention (CDC), the European Centre for Disease and Prevention (ECDC), German Robert Koch Institute, the American Dental Association, U.S. Occupational Safety and Health Administration, American Society of Dentist Anesthesiologists, British Association of Oral and Maxillofacial Surgeons, The French Society of Stomatology, Maxillo-Facial Surgery and Oral Surgery, British Society of Oral Surgeons etc.) recommending N95 /FFP2 also for non AGPs over the less expensive and more readily available medical masks (Bartoszko et al., 2020). Estimations in March 2020 for the U.S. indicated that only about 1% of the protective devices needed for the pandemic case had been stockpiled for HCWs (Berenson, 2020). Although, at least in most countries, this severe shortage of N95 respirators or FFP-2 masks or even medical masks experienced during the onset of the pandemic has eased and respirator stockpiles widely have been replenished by now due to widespread sessional use (Warnakulasuriya, 2020),resterilisation, increase in production output and lockdown spillover effects, the general use of N95/N99 respirators /FFP2 or 3 masks should be well considered and rather be reserved for high risk indications,(Bartoszko et al., 2020). According to Zimmermann &Nkenke, for routine care of low-risk patients (i.e. symptom free) the use of medical masks and gloves to protect against droplet transmission is considered sufficient (Zimmermann et al., 2020).

As a consequence, at least under pandemic conditions, to save resources (Zimmermann et al., 2020)and according to available evidence as presented in this paper, it may seem reasonable to differentiate between low and high risk dental and OMFS procedures, with just the latter ones requiring special precautions to prevent droplet and especially aerosolized disease transmission. For low risk treatments, current empirical data and the absence of clear scientific evidence for aerosol transmission of SARS-CoV-2 provide sufficient rationale for the use of surgical masks (Sommerstein et al., 2020), which should apply in analogy for in dentistry and OMFS, as well. As basic methods, e.g. during surgical procedures, Ge et al.(Ge et al., 2020) recommended to apply preprocedural mouth rinse and to put the patient in a supine position to avoid working in the breath way of the patient. Next, filtering of potentially contaminated aerosols can be achieved either by chairside high volume evacuators (HVE) or more expensive HEPA (high efficiency particulate arrestor) filters. HVE filters reduce contamination by the operating site by around 90% (Narayana et al., 2016), HEPA filters can remove 99.7% of particles measuring 0.3μm in diameter(Ge et al., 2020). In addition, even the use of negative pressure for ORs has been claimed mandatory to prevent spread of aerosols(Zimmermann et al., 2020). As such devices may be supposed to be frequently lacking in outpatient surgery settings, and as poor air ventilation prolongs the amount of time that aerosols remain airborne (Klompas et al., 2020),the use of the operating room should be discontinued for at least 15 minutes after the patient has left to allow for settling of aerosols and adequate room ventilation (Yao et al., 2020)to reduce aerosols before cleaning and surface disinfection starts (Zimmermann et al., 2020).Further options in OMFS surgery are e.g. the use of osteotomes whenever possible, of self-drilling screws and application of electric cautery using lowest possible power and a smoke evacuation system(Zimmermann et al., 2020).

## SUMMARY

In conclusion, According to available very weak/inconclusive evidence, transmission of SARS-CoV-2 via infective aerosol during AGPS, so far, must remain speculative and controversial. As, however, this is a probable opportunistic way of transmission which at least cannot be sufficiently excluded and therefore should not be dismissed out of hand prematurely, proper and equally important properly applied protective equipment (i.e., N95 respirators or FFP-2 masks or above regarding mouth and nose protection) should always be used during AGPs. To stay on the side of caution, it is therefore mandatory that aerosols as possible SARS-CoV-2 transmission routes in dental and OMFS surgery must be recognized and efficiently controlled, even more so when dealing with patients with suspected or confirmed COVID-19 in order to prevent spreading not only among HCWs, but also to avoid cross-infection between patients especially in out-patient settings with poor ventilation and air filtering. Further studies to clarify the role of aerosols vs droplets in OMFS, however, are clearly needed. Last but not least, there is urgent need for studies comparing respirators to surgical masks during dental and OMFS AGPs for protection against COVID-19 transmission.

## Data Availability

its systematic review, not required

---

**Supplemental file 1.** PICOTS database search strategy.

| Database | Search (August 28, 2020) |
| --- | --- |
| **PubMed** | (coronavirus OR COVID-19 OR acute respiratory distress syndrome OR SARS-CoV-2 OR Severe acute respiratory syndrome coronavirus 2 OR coronavirus disease OR novel coronavirus OR Coronavirus disease-19 OR Severe Acute Respiratory Syndrome Coronavirus 2 OR 2019-nCoV) AND (dental OR oral surgery OR maxillofacial surgery OR dental practice) AND (aerosol generating procedures OR bioaerosols OR airborne OR droplet OR aerosolization) AND (PPE OR surgical mask OR personal protective equipment OR N95 OR FFP OR respirators OR powered air purified respirator OR surgical mask) AND (randomized clinical trial OR clinical study OR in vitro study OR cadaveric study) |
| **Scopus** | (coronavirus OR COVID-19 OR acute respiratory distress syndrome OR SARS-CoV-2 OR Severe acute respiratory syndrome coronavirus 2 OR coronavirus disease OR novel coronavirus OR Coronavirus disease-19 OR Severe Acute Respiratory Syndrome Coronavirus 2 OR 2019-nCoV) AND (dental OR oral surgery OR maxillofacial surgery OR dental practice) AND (aerosol generating procedures OR bioaerosols OR airborne OR droplet OR aerosolization) AND (PPE OR personal protective equipment OR N95 OR FFP OR respirators OR powered air purified respirator OR surgical mask) AND (randomized clinical trial OR clinical study) |
| **Cochrane** | (coronavirus OR COVID-19 OR acute respiratory distress syndrome OR SARS-CoV-2 OR Severe acute respiratory syndrome coronavirus 2 OR coronavirus disease OR novel coronavirus OR Coronavirus disease-19 OR Severe Acute Respiratory Syndrome Coronavirus 2 OR 2019-nCoV) AND (dental OR oral surgery OR maxillofacial surgery OR dental practice) AND (aerosol generating procedures OR bioaerosols OR airborne OR droplet OR aerosolization) AND (PPE OR personal protective equipment OR N95 OR FFP OR respirators OR powered air purified respirator OR surgical mask) AND (randomized clinical trial OR clinical study) |

## References

2020 W: World Helath Organization: Rational use of of personal protective equipment (PPE) for coronavirus disease /Covid-19): interim guidance 19 March 2020. Geneva, World Health Organization. 2020.

Ai ZT, Huang T, Melikov AK: Airborne transmission of exhaled droplet nuclei between occupants in a room with horizontal air distribution. Building and Environment 163:106328, 2019.

Al-Eid RA, Ramalingam S, Sundar C, Aldawsari M, Nooh N: Detection of Visually Imperceptible Blood Contamination in the Oral Surgical Clinic using Forensic Luminol Blood Detection Agent. Journal of International Society of Preventive & Community Dentistry 8:327–332, 2018.

al CDe: Clinical Infectoius Diseases ciaa

Backer JA, Klinkenberg D, Wallinga J: Incubation period of 2019 novel coronavirus (2019-nCoV) infections among travellers from Wuhan, China, 20-28 January 2020. Euro Surveill 25:2020.

Balazy A, Toivola M, Adhikari A, Sivasubramani SK, Reponen T, Grinshpun SA: Do N95 respirators provide 95% protection level against airborne viruses, and how adequate are surgical masks? Am J Infect Control 34:51-57, 2006.

BAOMS: BAOMS –Guidance for the care of OMFSand Oral Surgerypatients where COVID is prevalent. 2020.

Bartoszko JJ, Farooqi MAM, Alhazzani W, Loeb M: Medical masks vs N95 respirators for preventing COVID-19 in healthcare workers: A systematic review and meta-analysis of randomized trials. Influenza Other Respir Viruses 14:365–373, 2020.

Bennett AM, Fulford MR, Walker JT, Bradshaw DJ, Martin MV, Marsh PD: Microbial aerosols in general dental practice. Br Dent J 189:664–667, 2000.

Bentley CD, Burkhart NW, Crawford JJ: Evaluating spatter and aerosol contamination during dental procedures. Journal of the American Dental Association (1939) 125:579–584, 1994.

Berenson T: States need medical supplies to fight coronavirus. Can the National Stockpile keep up with the demand? 2020.

Bin SY, Heo JY, Song MS, Lee J, Kim EH, Park SJ, Kwon HI, Kim SM, Kim YI, Si YJ, Lee IW, Baek YH, Choi WS, Min J, Jeong HW, Choi YK: Environmental Contamination and Viral Shedding in MERS Patients During MERS-CoV Outbreak in South Korea. Clin Infect Dis 62:755–760, 2016.

Burdorf A, Porru F, Rugulies R: The COVID-19 (Coronavirus) pandemic: consequences for occupational health. Scand J Work Environ Health 46:229–230, 2020.

Centers for Disease Control and Prevention: Guidance for Dental Settings : Interim Infection Prevention and Control Guidance for Dental Settings During the COVID-19 Response. Centers for Disease Control and Prevention. 2020a.

Centers for Disease Control and Prevention: Interim Infection Prevention and Control Recommendations for Patients with Suspected or Confirmed Coronavirus Disease 2019 (COVID- 19) in Healthcare Settings March 10, 2020. 2020b.

Chen J: Pathogenicity and transmissibility of 2019-nCoV-A quick overview and comparison with other emerging viruses. Microbes Infect 22:69–71, 2020.

Chen J, Qi T, Liu L, Ling Y, Qian Z, Li T, Li F, Xu Q, Zhang Y, Xu S, Song Z, Zeng Y, Shen Y, Shi Y, Zhu T, Lu H: Clinical progression of patients with COVID-19 in Shanghai, China. J Infect 80:e1–e6, 2020.

Chu DK, Akl EA, Duda S, Solo K, Yaacoub S, Schunemann HJ, authors C-SURGEs: Physical distancing, face masks, and eye protection to prevent person-to-person transmission of SARS-CoV-2 and COVID-19: a systematic review and meta-analysis. Lancet 395:1973–1987, 2020.

Douedi S DH: Precautions, Bloodborne, Contact, and Droplet. StatPearls Publishing.

Earnest R, Loesche W: Measuring harmful levels of bacteria in dental aerosols. Journal of the American Dental Association (1939) 122:55–57, 1991.

England. PH: COVID-19: infection prevention and control guidance. 2020.

Essam Ahmed Al-Moraissi NC: Are the beneficial effects of occlusal splint therapy beyond the placebo effect in managing temporomandibular disorders? PROSPERO 2020.

European Centre for Disease Prevention and Control: Guidance for wearing and removing personal protective equipment in healthcare settings for the care of patients with suspected or confirmed COVID-19. European Centre for Disease Prevention and Control. 2020.

French Society of Stomatology M-FS, Oral S: Practitioners specialized in oral health and coronavirus disease 2019: Professional guidelines from the French society of stomatology, maxillofacial surgery and oral surgery, to form a common front against the infectious risk. J Stomatol Oral Maxillofac Surg 121:155–158, 2020a.

French Society of Stomatology M-FS, Oral S: Practitioners specialized in oral health and coronavirus disease 2019: Professional guidelines from the French society of stomatology, maxillofacial surgery and oral surgery, to form a common front against the infectious risk. Journal of stomatology, oral and maxillofacial surgery 121:155–158, 2020b.

Frieden TR, Lee CT: Identifying and Interrupting Superspreading Events-Implications for Control of Severe Acute Respiratory Syndrome Coronavirus 2. Emerg Infect Dis 26:1059–1066, 2020.

Garcia Godoy LR, Jones AE, Anderson TN, Fisher CL, Seeley KML, Beeson EA, Zane HK, Peterson JW, Sullivan PD: Facial protection for healthcare workers during pandemics: a scoping review. BMJ Glob Health 5:2020.

Ge ZY, Yang LM, Xia JJ, Fu XH, Zhang YZ: Possible aerosol transmission of COVID-19 and special precautions in dentistry. J Zhejiang Univ Sci B 21:361–368, 2020.

Group C-DSERCW: Recommendations for the re-opening of dental services: a rapid review of international sources Version 1.3. 2020.

Guan WJ, Ni ZY, Hu Y, Liang WH, Ou CQ, He JX, Liu L, Shan H, Lei CL, Hui DSC, D. B, Li LJ, Zeng G, Yuen KY, Chen RC, Tang CL, Wang T, Chen PY, Xiang J, Li SY, Wang JL, Liang ZJ, Peng YX, Wei L, Liu Y, Hu YH, Peng P, Wang JM, Liu JY, Chen Z, Li G, Zheng ZJ, Qiu SQ, Luo J, Ye CJ, Zhu SY, Zhong NS, China Medical Treatment Expert Group for C: Clinical Characteristics of Coronavirus Disease 2019 in China. N Engl J Med 382:1708–1720, 2020.

H. F: Rapid expert consultation on the possibility of bioaerosol spread of SARS-CoV-2 for the COVID-19 pandemic. National Academies Press (US) (April 1, 2020).

Hamner L DP, Capron I, et al: igh SARS-CoV-2 Attack Rate Following Exposure at a Choir Practice — Skagit County, Washington. MMWR Morb Mortal Wkly Rep March 2020:606–610, 2020.

Harrel SK: Airborne spread of disease--the implications for dentistry. Journal of the California Dental Association 32:901–906, 2004.

Harrel SK, Molinari J: Aerosols and splatter in dentistry: a brief review of the literature and infection control implications. Journal of the American Dental Association (1939) 135:429–437, 2004.

Health and Safety Executive: Evaluating the protection afforded by surgical masks against influenza bioaerosols: Gross protection of surgical masks compared to filtering facepiece respirators. 2008.

Huang C, Wang Y, Li X, Ren L, Zhao J, Hu Y, Zhang L, Fan G, Xu J, Gu X, Cheng Z, Yu T, Xia J, Wei Y, Wu W, Xie X, Yin W, Li H, Liu M, Xiao Y, Gao H, Guo L, Xie J, Wang G, Jiang R, Gao Z, Jin Q, Wang J, Cao B: Clinical features of patients infected with 2019 novel coronavirus in Wuhan, China. Lancet 395:497–506, 2020.

Ishihama K, Iida S, Koizumi H, Wada T, Adachi T, Isomura-Tanaka E, Yamanishi T, Enomoto A, Kogo M: High incidence of blood exposure due to imperceptible contaminated splatters during oral surgery. Journal of oral and maxillofacial surgery : official journal of the American Association of Oral and Maxillofacial Surgeons 66:704–710, 2008.

Ishihama K, Koizumi H, Wada T, Iida S, Tanaka S, Yamanishi T, Enomoto A, Kogo M: Evidence of aerosolised floating blood mist during oral surgery. J Hosp Infect 71:359–364, 2009.

Klompas M, Baker MA, Rhee C: Airborne Transmission of SARS-CoV-2. Jama 324:441–442, 2020.

Korth J, Wilde B, Dolff S, Anastasiou OE, Krawczyk A, Jahn M, Cordes S, Ross B, Esser S, Lindemann M, Kribben A, Dittmer U, Witzke O, Herrmann A: SARS-CoV-2-specific antibody detection in healthcare workers in Germany with direct contact to COVID-19 patients. J Clin Virol 128:104437, 2020.

Kutter JS, Spronken MI, Fraaij PL, Fouchier RA, Herfst S: Transmission routes of respiratory viruses among humans. Curr Opin Virol 28:142–151, 2018.

Li RW, Leung KW, Sun FC, Samaranayake LP: Severe acute respiratory syndrome (SARS) and the GDP. Part II: implications for GDPs. Br Dent J 197:130–134, 2004.

Liu L, Wei Q, Alvarez X, Wang H, Du Y, Zhu H, Jiang H, Zhou J, Lam P, Zhang L, Lackner A, Qin C, Chen Z: Epithelial cells lining salivary gland ducts are early target cells of severe acute respiratory syndrome coronavirus infection in the upper respiratory tracts of rhesus macaques. J Virol 85:4025–4030, 2011.

Liu Y, Ning Z, Chen Y, Guo M, Liu Y, Gali NK, Sun L, Duan Y, Cai J, Westerdahl D, Liu X, Xu K, Ho KF, Kan H, Fu Q, Lan K: Aerodynamic analysis of SARS-CoV-2 in two Wuhan hospitals. Nature 582:557–560, 2020.

Loeb M, Dafoe N, Mahony J, John M, Sarabia A, Glavin V, Webby R, Smieja M, Earn DJ, Chong S, Webb A, Walter SD: Surgical mask vs N95 respirator for preventing influenza among health care workers: a randomized trial. JAMA 302:1865–1871, 2009.

Loh NW, Tan Y, Taculod J, Gorospe B, Teope AS, Somani J, Tan AYH: The impact of high-flow nasal cannula (HFNC) on coughing distance: implications on its use during the novel coronavirus disease outbreak. Can J Anaesth 67:893–894, 2020.

Long Y, Hu T, Liu L, Chen R, Guo Q, Yang L, Cheng Y, Huang J, Du L: Effectiveness of N95 respirators versus surgical masks against influenza: A systematic review and meta-analysis. J Evid Based Med 13:93–101, 2020.

Lu C-w, Liu X-f, Jia Z-f: 2019-nCoV transmission through the ocular surface must not be ignored. The Lancet 395:e39, 2020.

Marui VC, Souto MLS, Rovai ES, Romito GA, Chambrone L, Pannuti CM: Efficacy of preprocedural mouthrinses in the reduction of microorganisms in aerosol: A systematic review. Journal of the American Dental Association (1939) 150:1015–1026 e1011, 2019.

Meng L, Hua F, Bian Z: Coronavirus Disease 2019 (COVID-19): Emerging and Future Challenges for Dental and Oral Medicine. J Dent Res 99:481–487, 2020.

Micik RE, Miller RL, Leong AC: Studies on dental aerobiology. 3. Efficacy of surgical masks in protecting dental personnel from airborne bacterial particles. J Dent Res 50:626–630, 1971.

Micik RE, Miller RL, Mazzarella MA, Ryge G: Studies on dental aerobiology. I. Bacterial aerosols generated during dental procedures. J Dent Res 48:49–56, 1969.

Miller RL, Micik RE: Air pollution and its control in the dental office. Dent Clin North Am 22:453–476, 1978.

Moher D, Shamseer L, Clarke M, Ghersi D, Liberati A, Petticrew M, Shekelle P, Stewart LA, Group P-P: Preferred reporting items for systematic review and meta-analysis protocols (PRISMA-P) 2015 statement. Syst Rev 4:1, 2015.

Monje Gil F, Cebrián Carretero JL, López-Cedrún Cembranos JL, Redondo Alamillos M, Valdés Beltrán A, Almeida Parra F, Gómez García E, Díaz-Mauriñoy Garrido-Lestache JC, Tousidonis Rial MA, Ruiz-Laza L, Sastre Pérez J, Ranz Colio Á, Acebal Blanco F, Rubio-Palau J, Pedro Marina Md, Redondo González LM, Pla Esparza A, Infante-Cossío P: Manejo de pacientes en cirugía oral y maxilofacial durante el periodo de crisis y de control posterior de la pandemia de COVID-19. Revista Española de Cirugía Oral y Maxilofacial 42:51–59, 2020.

Morawska L, Milton DK: It is Time to Address Airborne Transmission of COVID-19. Clin Infect Dis 2020.

Narayana TV, Mohanty L, Sreenath G, Vidhyadhari P: Role of preprocedural rinse and high volume evacuator in reducing bacterial contamination in bioaerosols. J Oral Maxillofac Pathol 20:59–65, 2016.

Nogler M, Lass-Florl C, Ogon M, Mayr E, Bach C, Wimmer C: Environmental and body contamination through aerosols produced by high-speed cutters in lumbar spine surgery. Spine (Phila Pa 1976) 26:2156–2159, 2001a.

Nogler M, Lass-Florl C, Wimmer C, Bach C, Kaufmann C, Ogon M: Aerosols produced by high-speed cutters in cervical spine surgery: extent of environmental contamination. Eur Spine J 10:274–277, 2001b.

Nogler M, Lass-Florl C, Wimmer C, Mayr E, Bach C, Ogon M: Contamination during removal of cement in revision hip arthroplasty. A cadaver study using ultrasound and high-speed cutters. J Bone Joint Surg Br 85:436–439, 2003.

Occupational safety and health administrations: OSHA Guidance for Dentistry Workers and Employers. 2020.

Organization WH: Modes of transmission of virus causing COVID-19: Implications for IPC precaution recommendations. Geneva;. 2020.

Panesar K, Dodson T, Lynch J, Bryson-Cahn C, Chew L, Dillon J: Evolution of COVID-19 Guidelines for University of Washington Oral and Maxillofacial Surgery Patient Care. Journal of oral and maxillofacial surgery : official journal of the American Association of Oral and Maxillofacial Surgeons 78:1136–1146, 2020.

Patel ZM, Fernandez-Miranda J, Hwang PH, Nayak JV, Dodd RL, Sajjadi H, Jackler RK: In Reply: Precautions for Endoscopic Transnasal Skull Base Surgery During the COVID-19 Pandemic. Neurosurgery 87:E162–E163, 2020.

Paulo AC, Correia-Neves M, Domingos T, Murta AG, Pedrosa J: Influenza infectious dose may explain the high mortality of the second and third wave of 1918-1919 influenza pandemic. PLoS One 5:e11655, 2010.

Peditto M, Scapellato S, Marciano A, Costa P, Oteri G: Dentistry during the COVID-19 Epidemic: An Italian Workflow for the Management of Dental Practice. Int J Environ Res Public Health 17:2020.

Peng X, Xu X, Li Y, Cheng L, Zhou X, Ren B: Transmission routes of 2019-nCoV and controls in dental practice. Int J Oral Sci 12:9, 2020.

Pluim JME, Jimenez-Bou L, Gerretsen RRR, Loeve AJ: Aerosol production during autopsies: The risk of sawing in bone. Forensic Sci Int 289:260–267, 2018.

Public Health Agency of Canada: Coronavirus disease (COVID-19): For health professionals. Public Health Agency of Canada 2020.

Public Health England: When to use a surgical face mask or FFP3 respirator. 2020a.

Public Health England PHE: COVID-19: infection prevention and control guidance. 2020b.

Qian Y, Willeke K, Grinshpun SA, Donnelly J, Coffey CC: Performance of N95 respirators: filtration efficiency for airborne microbial and inert particles. Am Ind Hyg Assoc J 59:128–132, 1998.

Radonovich LJ, Jr., Simberkoff MS, Bessesen MT, Brown AC, Cummings DAT, Gaydos CA, Los JG, Krosche AE, Gibert CL, Gorse GJ, Nyquist AC, Reich NG, Rodriguez-Barradas MC, Price CS, Perl TM, Res Pi: N95 Respirators vs Medical Masks for Preventing Influenza Among Health Care Personnel: A Randomized Clinical Trial. JAMA 322:824–833, 2019.

Rautemaa R, Nordberg A, Wuolijoki-Saaristo K, Meurman JH: Bacterial aerosols in dental practice - a potential hospital infection problem? J Hosp Infect 64:76–81, 2006.

Remuzzi A, Remuzzi G: COVID-19 and Italy: what next? Lancet 395:1225–1228, 2020.

Rothe C, Schunk M, Sothmann P, Bretzel G, Froeschl G, Wallrauch C, Zimmer T, Thiel V, Janke C, Guggemos W, Seilmaier M, Drosten C, Vollmar P, Zwirglmaier K, Zange S, Wolfel R, Hoelscher M: Transmission of 2019-nCoV Infection from an Asymptomatic Contact in Germany. N Engl J Med 382:970–971, 2020.

Roy CJ, Milton DK: Airborne transmission of communicable infection--the elusive pathway. N Engl J Med 350:1710–1712, 2004.

Sacchetti R, Baldissarri A, De Luca G, Lucca P, Stampi S, Zanetti F: Microbial contamination in dental unit waterlines: comparison between Er:YAG laser and turbine lines. Ann Agric Environ Med 13:275–279, 2006.

Santarpia J, Rivera D, Herrera V, Morwitzer J, Creager H, Santarpia G, Crown K, Brett-Major D, Schnaubelt E, Broadhurst J, Lawler J, Reid SP, Lowe J: Transmission Potential of SARS-CoV-2 in Viral Shedding Observed at the University of Nebraska Medical Center. medRxiv. 2020a.

Santarpia JL, Rivera DN, Herrera V, Morwitzer MJ, Creager H, Santarpia GW, Crown KK, Brett-Major D, Schnaubelt E, Broadhurst MJ, Lawler JV, Reid SP, Lowe JJ: Transmission Potential of SARS-CoV-2 in Viral Shedding Observed at the University of Nebraska Medical Center. medRxiv 2020.2003.2023.20039446, 2020b.

Schmidt SB, Gruter L, Boltzmann M, Rollnik JD: Prevalence of serum IgG antibodies against SARS-CoV-2 among clinic staff. PLoS One 15:e0235417, 2020.

Schwartz KL, Murti M, Finkelstein M, Leis JA, Fitzgerald-Husek A, Bourns L, Meghani H, Saunders A, Allen V, Yaffe B: Lack of COVID-19 transmission on an international flight. CMAJ 192:E410, 2020.

Seto WH, Tsang D, Yung RW, Ching TY, Ng TK, Ho M, Ho LM, Peiris JS, Advisors of Expert SgoHA: Effectiveness of precautions against droplets and contact in prevention of nosocomial transmission of severe acute respiratory syndrome (SARS). Lancet 361:1519–1520, 2003.

Shiu EYC, Leung NHL, Cowling BJ: Controversy around airborne versus droplet transmission of respiratory viruses: implication for infection prevention. Curr Opin Infect Dis 32:372–379, 2019.

Singh A, Shiva Manjunath RG, Singla D, Bhattacharya HS, Sarkar A, Chandra N: Aerosol, a health hazard during ultrasonic scaling: A clinico-microbiological study. Indian journal of dental research : official publication of Indian Society for Dental Research 27:160–162, 2016.

Sommerstein R, Fux CA, Vuichard-Gysin D, Abbas M, Marschall J, Balmelli C, Troillet N, Harbarth S, Schlegel M, Widmer A, Balmelli C, Eisenring M-C, Harbarth S, Marschall J, Pittet D, Sax H, Schlegel M, Schweiger A, Senn L, Troillet N, Widmer AF, Zanetti G, Swissnoso: Risk of SARS-CoV-2 transmission by aerosols, the rational use of masks, and protection of healthcare workers from COVID-19. Antimicrobial Resistance & Infection Control 9:100, 2020.

Szymanska J, Sitkowska J: Bacterial contamination of dental unit waterlines. Environ Monit Assess 185:3603–3611, 2013.

Teleman MD, Boudville IC, Heng BH, Zhu D, Leo YS: Factors associated with transmission of severe acute respiratory syndrome among health-care workers in Singapore. Epidemiol Infect 132:797–803, 2004.

Thamboo A, Lea J, Sommer DD, Sowerby L, Abdalkhani A, Diamond C, Ham J, Heffernan A, Cai Long M, Phulka J, Wu YQ, Yeung P, Lammers M: Clinical evidence based review and recommendations of aerosol generating medical procedures in otolaryngology - head and neck surgery during the COVID-19 pandemic. J Otolaryngol Head Neck Surg 49:28, 2020.

The American Society of Dentist Anesthesiologists: Interim Guidance For Dentist Anesthesiologists Practicing In The Office-Based Setting During The COVID-19 Pandemic. 2020.

To KK, Tsang OT, Yip CC, Chan KH, Wu TC, Chan JM, Leung WS, Chik TS, Choi CY, Kandamby DH, Lung DC, Tam AR, Poon RW, Fung AY, Hung IF, Cheng VC, Chan JF, Yuen KY: Consistent Detection of 2019 Novel Coronavirus in Saliva. Clin Infect Dis 71:841–843, 2020.

Tran K, Cimon K, Severn M, Pessoa-Silva C, Conly J: Aerosol-generating procedures and risk of transmission of acute respiratory infections: a systematic review. CADTH Technol Overv 3:e3201, 2013.

Tran K, Cimon K, Severn M, Pessoa-Silva CL, Conly J: Aerosol generating procedures and risk of transmission of acute respiratory infections to healthcare workers: a systematic review. PLoS One 7:e35797, 2012.

van Doremalen N, Bushmaker T, Morris DH, Holbrook MG, Gamble A, Williamson BN, Tamin A, Harcourt JL, Thornburg NJ, Gerber SI, Lloyd-Smith JO, de Wit E, Munster VJ: Aerosol and Surface Stability of SARS-CoV-2 as Compared with SARS-CoV-1. N Engl J Med 382:1564–1567, 2020.

Veena HR, Mahantesha S, Joseph PA, Patil SR, Patil SH: Dissemination of aerosol and splatter during ultrasonic scaling: a pilot study. J Infect Public Health 8:260–265, 2015.

Wang J, Zhou M, Liu F: Reasons for healthcare workers becoming infected with novel coronavirus disease 2019 (COVID-19) in China. J Hosp Infect 105:100–101, 2020.

Warnakulasuriya S: Protecting dental manpower from COVID-19 infection. Oral Dis 2020.

Wax RS, Christian MD: Practical recommendations for critical care and anesthesiology teams caring for novel coronavirus (2019-nCoV) patients. Can J Anaesth 67:568–576, 2020.

Wen Z, Yu L, Yang W, Hu L, Li N, Wang J, Li J, Lu J, Dong X, Yin Z, Zhang K: Assessment the protection performance of different level personal respiratory protection masks against viral aerosol. Aerobiologia (Bologna) 29:365–372, 2013.

World Health Organization: Rational use of personal protective equipment for coronavirus disease 2019 (COVID-19). World Health Organization 2020.

Wu Z, McGoogan JM: Characteristics of and Important Lessons From the Coronavirus Disease 2019 (COVID-19) Outbreak in China: Summary of a Report of 72314 Cases From the Chinese Center for Disease Control and Prevention. JAMA 323:1239–1242, 2020.

Xu H, Zhong L, Deng J, Peng J, Dan H, Zeng X, Li T, Chen Q: High expression of ACE2 receptor of 2019-nCoV on the epithelial cells of oral mucosa. Int J Oral Sci 12:8, 2020.

Yamada H, Ishihama K, Yasuda K, Hasumi-Nakayama Y, Shimoji S, Furusawa K: Aerial dispersal of blood-contaminated aerosols during dental procedures. Quintessence Int 42:399–405, 2011.

Yao M, Zhang L, Ma J, Zhou L: On airborne transmission and control of SARS-Cov-2. Sci Total Environ 731:139178, 2020.

Zemouri C, de Soet H, Crielaard W, Laheij A: A scoping review on bio-aerosols in healthcare and the dental environment. PLoS One 12:e0178007, 2017.

Zimmermann M, Nkenke E: Approaches to the management of patients in oral and maxillofacial surgery during COVID-19 pandemic. Journal of cranio-maxillo-facial surgery : official publication of the European Association for Cranio-Maxillo-Facial Surgery 48:521–526, 2020.

